# A Bayesian model for repeated cross-sectional epidemic prevalence survey data

**DOI:** 10.1101/2025.04.16.25325936

**Authors:** Nicholas Steyn, Marc Chadeau-Hyam, Paul Elliott, Christl A. Donnelly

## Abstract

Epidemic prevalence surveys monitor the spread of an infectious disease by regularly testing representative samples of a population for infection. State-of-the-art Bayesian approaches for analysing epidemic survey data were constructed independently and under pressure during the COVID-19 pandemic. In this paper, we compare two existing approaches (one leveraging Bayesian P-splines and the other approximate Gaussian processes) with a novel approach (leveraging a random walk and fit using sequential Monte Carlo) for smoothing and performing inference on epidemic survey data. We use our simpler approach to investigate the impact of survey design and underlying epidemic dynamics on the quality of estimates. We then incorporate these considerations into the existing approaches and compare all three on simulated data and on real-world data from the SARS-CoV-2 REACT-1 prevalence study in England. All three approaches, once appropriate considerations are made, produce similar estimates of infection prevalence; however, estimates of the growth rate and instantaneous reproduction number are more sensitive to underlying assumptions. Interactive notebooks applying all three approaches are also provided alongside recommendations on hyperparameter selection and other practical guidance, with some cases resulting in orders-of-magnitude faster runtime.

**Author summary:** Understanding how infections spread in a population is crucial during an epidemic, and largescale surveys that test people for current infection can provide valuable insights. These surveys are resource-intensive and the data they produce can be noisy and hard to interpret. In this study, we investigate how three different statistical approaches (two established and one novel) can explore such data. We found that some common modelling choices, particularly the treatment of observation noise, can meaningfully shape results. Our findings highlight the need for careful, robust methods to help researchers and public health officials make best use of existing data, design more effective surveys, and extract clearer insights from future studies.

## 1 Introduction

The aims of infectious disease surveillance include describing disease burden, monitoring trends, and identifying outbreaks and novel pathogens through the collection, analysis, and interpretation of health data [1]. A range of passive systems, such as automated reporting from healthcare facilities, and active systems, such as field investigations, are used to gather these data. Recently, novel surveillance methods, such as wastewater testing [2], mobile phone data [3, 4], and social media monitoring [5], have been developed to complement traditional surveillance methods. While these methods provide an unprecedented volume of data, they are often subject to biases and limitations that make reliable inference and interpretation difficult [6].

Large-scale prevalence surveys are another tool for the surveillance of infectious diseases. These surveys typically use random sampling methods to produce estimates of the prevalence of infection in a given population. The quality of data collected also allows for the robust estimation of epidemiological quantities such as prevalence *P*_*t*_, the growth rate *r*_*t*_, and the instantaneous reproduction number *R*_*t*_ [7]. Established during the COVID-19 pandemic, the REeal-time Assessment of Community Transmission (REACT-1) study in England [8] processed over 2.5 million self-administered throat and nose samples between May 2020 and March 2022. The ONS Coronavirus Infection Survey [9] monitored the spread of SARS-CoV-2 in the United Kingdom, also processing millions of samples over the course of the pandemic. These surveys have been instrumental in understanding, for example, the spread of SARS-CoV-2 [8], the dynamics of infection hospitalisation and infection fatality ratios [10, 11], and the impact of vaccination [12].

Implementing such large-scale surveys is expensive; for example, the ONS Coronavirus Infection Survey cost £988.5 million (until September 2023) [13]. It is therefore critical to maximise the information extracted from the data — not only to justify cost, but also to support timely decision-making and improve policy relevance. This is made challenging by the substantial noise inherent in point prevalence estimates. Even with an ideal survey design, large sample size, and only ignorable non-response, the coefficient of variation of common binomial proportion estimators can be large [14], particularly when infection prevalence is low.

Even when the goal is to perform “model-free” inference (making inferences that reflect only the data, rather than modelling assumptions), noise in these data often necessitates the use of smoothing methods to improve the quality of the outputs. Smoothing methods impose constraints on day-to-day variability, allowing data from multiple days to inform point estimates. Smoothing allows the researcher to make more confident statements, increasing information yield or reducing the sample size needed for a given confidence level. However, all smoothing methods make assumptions about the underlying process, whether explicitly or implicitly. Even seemingly simple smoothing methods can introduce substantial bias, turning “model-light” estimates (that rely only minimally on modelling assumptions) into “model-heavy” estimates (that are strongly shaped by modelling assumptions) that may not accurately reflect reality.

Here we introduce a novel Bayesian approach for smoothing and performing inference on epidemic survey data, referred to as SIMPLE (Survey Inference Method for Prevalence and other Latent variables in Epidemiology). This approach is based on hidden-state models and sequential Monte Carlo (SMC) methods [15, 16], and is designed to be flexible enough to incorporate key assumptions of existing approaches, while avoiding some of the computational and mathematical complexity of these approaches. We compare the SIMPLE approach to two existing approaches, one by Eales, Ainslie, Walters, et al. [17] that leverages Bayesian P-splines and another by Abbott and Funk [7] that uses Gaussian processes approximations. We refer to these approaches as the Eales approach and the Abbott approach respectively. All three approaches are presented using common and general notation, allowing for direct comparison of their assumptions, performance, and results.

The results of this paper are structured in three parts. First, we use the SIMPLE approach to investigate the impact of survey design (such as the number of samples and individual response bias) and underlying epidemic dynamics (such as the variability in the growth rate) on the quality of estimates, highlighting key modelling decisions that should be made when analysing epidemic survey data. Second, we demonstrate how the approaches of Eales and Abbott can be adapted to account for these considerations, and compare all three (improved) approaches on simulated data to highlight similarities and differences in their performance. The use of simulated data provides a ground truth that allows us to calculate and compare quantities such as statistical coverage (the percentage of times that credible intervals of a given level contain the true value). Third, we apply all three approaches to real-world data from the REACT-1 study, covering the COVID-19 pandemic between May 2020 and March 2022 in England, comparing the quality of estimates and computational requirements of each approach. We provide recommendations for future modelling and survey design based on our findings.

Our goal is to provide a framework for understanding the impact of modelling decisions on the quality of estimates, to consolidate and improve state-of-the-art methods, and to make recommendations for future modelling and survey design. We also provide documented source code for all three approaches, notebooks demonstrating their use, and recommendations for hyperparameter selection and other practical considerations.

## 2 Materials and Methods

### 2.1 The SIMPLE approach

We introduce a suite of state-space models for simultaneously smoothing and performing inference on epidemic survey data. Each state-space model consists of an explicitly defined epidemic and observation model. The epidemic model encodes our assumptions about the unobserved dynamics of the epidemic, while the observation model describes how the observed data are generated from the underlying epidemic.

Two epidemic models are considered: one for estimating the daily exponential growth rate *r*_*t*_ in prevalence *P*_*t*_ and one for estimating the instantaneous reproduction number *R*_*t*_, infection incidence *I*_*t*_ and prevalence *P*_*t*_. Three observation models are considered: a basic model that assumes the number of positive swabs (diagnostic tests) follows a binomial distribution (as used by the original Eales approach), an extra-binomial model that accounts for overdispersion in the data, and a weighted model that accounts for survey weights.

We refer to the unknown time-varying quantities of interest, such as *r*_*t*_ and *P*_*t*_, as hidden states. Other unknown parameters that are not time-varying, such as the level of overdispersion in the extra-binomial model, are referred to as static parameters.

Throughout this paper, we use the term *prevalence* to describe the proportion of the population that would test positive. This quantity may differ from the proportion of the population with an active infection (due to imperfect test sensitivity/specificity) and from the proportion who are infectious (as test positivity can outlast infectiousness).

#### 2.1.1 Growth rate epidemic model

This model assumes that the daily growth rate *r*_*t*_ in prevalence follows a Gaussian random walk, encoding an assumption that *r*_*t*_ varies smoothly over time. Prevalence *P*_*t*_ on day *t* ∈ {1, 2, …, *T* } then varies exponentially with rate *r*_*t*_.

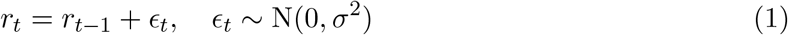

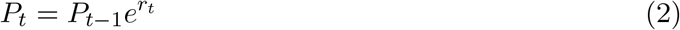

Coupled with initial distributions for *r*_0_ and *P*_0_, and a prior distribution for *σ*, equations 1 and 2 define the entire epidemic model. Parameter *σ* controls the smoothness of our estimates and is inferred from the data.

We use default initial distributions of *r*_0_ ∼ Uniform(−0.3, 0.3) and 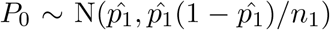, where 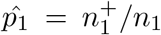 (the Wald interval), and default prior distribution *σ* ∼ Uniform(0, 0.2), although inferences are insensitive to the choice of these prior distributions (Supplementary Section 1).

As this model makes minimal assumptions about the underlying epidemic, we use it as the default epidemic model unless otherwise stated.

#### 2.1.2 Reproduction number epidemic model

Alternatively, we may want to estimate the instantaneous reproduction number *R*_*t*_, a popular alternative measure to *r*_*t*_ for characterising the rate of epidemic spread [18]. *R*_*t*_ is the average number of secondary infections generated by a primary infection at time *t* if an individual were to undergo their entire infectious period at this time. As with *r*_*t*_, we employ a Gaussian random walk to smooth estimates, now on log *R*_*t*_ to ensure positivity:

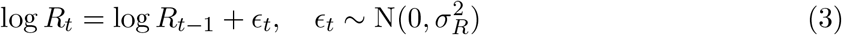

We then employ the renewal model [16, 19], which relates past infection incidence *I*_1:*t−*1_ to current infection incidence *I*_*t*_ through the instantaneous reproduction number *R*_*t*_ and a generation time distribution (representing the time from infection of an infector and their infectee, described by probability mass function *w*_*u*_):

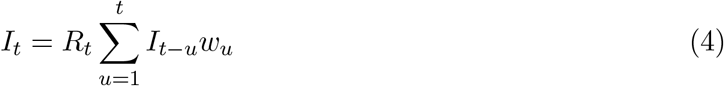

Finally, we relate prevalence *P*_*t*_ to infection incidence *I*_*t*_ through a test-sensitivity function *h*_*u*_ that describes how likely an individual is to test positive *u* days after infection. We do not consider imperfect test specificity, although this could be included with the addition of a constant term to *P*_*t*_:

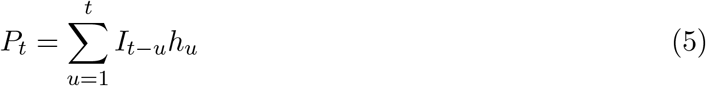

When applying the reproduction number epidemic model to data from the REACT-1 study, the generation time is assumed to follow a gamma distribution with mean 6.4 days and standard deviation 4.2 days [7], discretised by evaluating the gamma density function at integer time steps and normalising so Σ*w*_*u*_ = 1, and chosen to be consistent with the generation time distribution used in the published Abbott approach. There is evidence that the generation time of SARS-CoV-2 has shortened with more recent variants [20], and this should be kept in mind when interpreting our real-world results. In the absence of a REACT-1-specific test-sensitivity function, we use the pointwise central estimate of *h*_*u*_ from [21], reflecting the type of test used in REACT-1 (reverse transcription polymerase chain reaction (RT-PCR)), but not other study-specific factors that may impact sensitivity. This is the same mean test-sensitivity function as used in the Abbott approach.

Estimating *R*_*t*_ requires many additional assumptions about the underlying epidemic, and thus has more potential to bias estimates of *P*_*t*_. We focus on the growth rate epidemic model for much of this paper, although demonstrate the *R*_*t*_ estimator on real-world REACT-1 data.

#### 2.1.3 Basic observation model

Given *n*_*t*_ swabs conducted on day *t*, the observed number of positive swabs 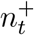 is modelled as a binomial random variable with probability *P*_*t*_:

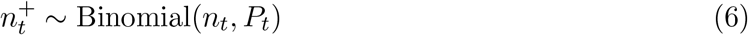

This is also the observation model used in the Eales approach as originally published.

#### 2.1.4 Extra-binomial observation model

Prevalence data are often overdispersed relative to the binomial distribution, exhibiting what is known as extra-binomial variation [22]. If we assume that the “observable” prevalence at time *t* is beta-distributed with mean equal to prevalence *P*_*t*_ and variance *ρP*_*t*_(1 − *P*_*t*_), our observation distribution becomes:

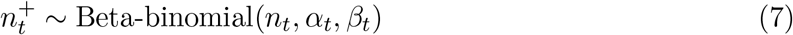

where 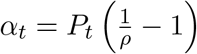 and 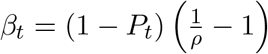.

The additional parameter *ρ* ∈ (0, 1) controls the modelled level of overdispersion in the data. Larger values of *ρ* indicate greater levels of overdispersion while letting *ρ* → 0 recovers the binomial model. By default, we use a Uniform(0, 0.01) prior distribution for this parameter. For context, the greatest upper bound of any 95% credible interval for *ρ* estimated from a real-world dataset is 0.0006.

This is our default observation model and is used in all subsequent analyses unless otherwise stated. It is common in other epidemiological settings to model overdispersed count data using the negative-binomial distribution [23], which is useful when the data have no natural upper limit. However, as the number of positive swabs are bounded above by the total number of swabs, we do not consider this here.

#### 2.1.5 Weighted observation model

Survey weights are used to account for bias in survey data arising from unequal probabilities of selection and/or response. Letting *w*_*t,i*_ be the (normalised) weight assigned to the *i*^*th*^ sample taken at time *t*, and *x*_*t,i*_ be the corresponding result (where *x*_*t,i*_ = 1 is individual *i* on day *t* tests positive, and zero otherwise), the weighted swab positivity is:

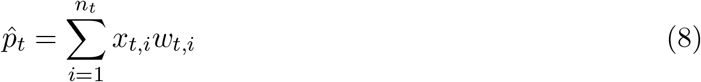

If the weights are uncorrelated with swab positivity, then 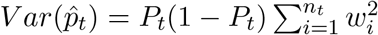. However, if the weights are correlated with the individual probability of testing positive, the variance of 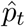 may be over or underestimated by this expression. The difference depends on the specific relationship between weights and outcome, which is generally unknown. Thus, we model 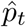 using a normal distribution with mean *P*_*t*_ and variance 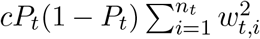, where *c* is an estimated scalar parameter:

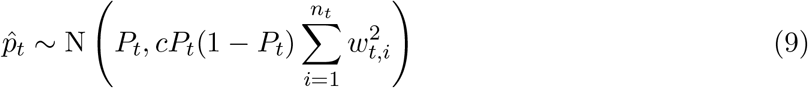

The observation distribution is no longer exact, in that we are approximating the unknown distribution of 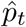 with a normal distribution. This is similar to the approximation used in the Abbott approach. By default, we use a Uniform(0.1, 10) prior distribution for *c*.

#### 2.1.6 Sequential Monte Carlo (SMC)

We use an SMC algorithm (also known as a particle filter), the bootstrap filter, to estimate the posterior distributions of the hidden states (such as *r*_*t*_, *P*_*t*_, and *R*_*t*_) at each time step *t*, given the observed data [16]. We also use this algorithm to estimate the posterior predictive distribution of 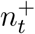, which is similar to the frequentist’s predictive distribution, and can be used to assess the quality of fit of the model when fitting to real-world data. Particle marginal Metropolis Hastings (PMMH) is used to estimate the static parameters, the uncertainty of which is then marginalised over in the final inference.

We run three chains of the PMMH algorithm, each for 100 iterations at a time, until the maximum Gelman-Rubin diagnostic 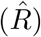 [24] is less than 1.05 and the minimum effective sample size (ESS) is greater than 100. These algorithms are detailed in full in Supplementary Section 2. We make the full source code (written natively in Julia [25]) available on GitHub, along with notebooks, to facilitate the application of these approaches to other datasets.

When fitting the model for the reproduction number, an additional “wind-in” period (default 10 days) is required to account for infection history prior to the first observation. After sampling from the standard prior distribution for *P*_0_, assumed values of *I*_*−*9:0_ are set to *P*_0_*/* Σ*h*_*u*_, ensuring the distribution of implied prevalence at time 0 is consistent with the chosen prior distribution. As 10 days may truncate the generation time distribution and test-sensitivity function, we rescale the generation time distribution to sum to 1 and the test-sensitivity function to sum to its original total over the combined length of past data and the wind-in period.

### 2.2 The Eales approach

The Eales approach [17] models logit-transformed *P*_*t*_ using Bayesian p-splines:

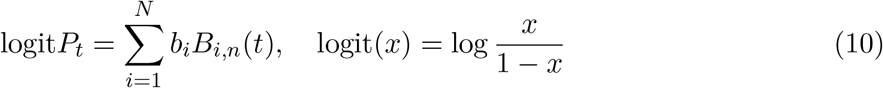

where *B*_*i,n*_(*t*) are basis functions and *b*_*i*_ are estimated spline coefficients. The coefficients give the value of the spline at the corresponding “knots” *t*_*i*_ while the basis functions allow for interpolation. By default, fourth-order basis functions *B*_*i*,4_(*t*) are used. We refer the reader to the original paper for the full construction [17].

The smoothness of these splines is controlled by the spacing of the spline knots (set by the user), and a second-order Gaussian random walk prior distribution on the spline coefficients, with standard deviation *σ*_*Eales*_ (estimated from the data):

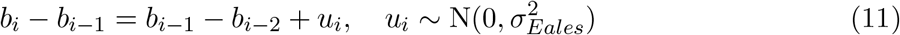

As logit*P*_*t*_ − logit*P*_*t−*1_ ≈ log *P*_*t*_ − log *P*_*t−*1_ = *r*_*t*_ for small *P*_*t*_, this is approximately equivalent to modelling the growth rate using a Gaussian random walk, a similar smoothing assumption to our growth rate epidemic model. This approximation can be improved by noting that logit*P*_*t*_ − logit*P*_*t−*1_ ≈ log *P*_*t*_ − log *P*_*t−*1_ + (*P*_*t*_ − *P*_*t−*1_) and the smoothness of *P*_*t*_ implies *P*_*t*_ − *P*_*t−*1_ is small.

By default, spline knots are placed approximately every 5 days (exactly 5 days when the duration of the data is divisible by 5) to balance flexibility with computational efficiency. As we generally work in integer time, there is no benefit to knot-spacing shorter than 1 day. If knots are placed on each day, we only need to evaluate the splines at the knot locations, and thus can work with *b*_*i*_ directly (i.e., we do not need to employ any splines). In this case, except for modelling the change in logit*P*_*t*_ instead of log *P*_*t*_, the model is equivalent to the SIMPLE (extra-binomial) model. We examine this equivalence further in Supplementary Section 3.

The original Eales approach modelled positive swabs 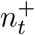 with a binomial distribution, equivalent to our basic observation model. We update this to use a beta-binomial distribution with overdispersion parameter *ρ*, equivalent to our extra-binomial observation model, improving both convergence of the algorithm and the quality-of-fit of the model (Supplementary Sections 3 and 4). We do not provide a weighted implementation of the Eales approach, although this could be achieved by using the weighted observation model described above.

Growth rates (in prevalence) are back-calculated from the fitted splines by setting *r*_*t*_ = log(*P*_*t*_*/P*_*t−*1_), where *P*_1:*T*_ are sampled from the posterior distribution of the splines. A weakly informative inverse-gamma prior distribution (*α* = *β* = 0.0001) is used for *σ*_*Eales*_, a uniform prior distribution is used for *ρ*, and a uniform prior distribution is used for the first two spline coefficients.

Rather than explicitly model incidence, the Eales approach makes a series of simplifying as-sumptions when estimating the reproduction number. Specifically, it assumes that at each independent time step *t, R*_*t*_ has been fixed for the past *τ* days (a trailing window approach akin to that used in EpiEstim [19]). If *τ* is chosen to be larger than both the maximum generation time and maximum duration of swab positivity, then the renewal equation (4) can be used to estimate *R*_*t*_ directly. In practice, *τ* = 14 days is used to prevent oversmoothing of *R*_*t*_, despite this likely being less than the maximum duration of test sensitivity for SARS-CoV-2 (which, according to [21], remains above 10% even after 21 days after infection). The trailing window approach is known to produce biased estimates of *R*_*t*_ [26], which can be partially accounted for by reporting estimates shifted by *τ/*2 days [27].

Eales, Ainslie, Walters, et al. [17] provide source code to fit this model using the R programming language [28] and Rstan. We adapt their code to fit the model using the actively developed interface to Stan, cmdstanr [29]. As with Rstan, inference is performed using Hamiltonian Monte Carlo (HMC) with the adaptive No-U-Turn Sampler (NUTS). We also adjust the hyperparameters of their algorithm (increasing the maximum tree-depth to 15 and decreasing the number of warm-up/sampling iterations to 200/300) to substantially improve convergence times, from at least 30 hours on the full REACT-1 dataset to less than 1 hour (Supplementary Section 4).

As in the SIMPLE approach, convergence is measured by running three chains and checking that maximum 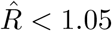 and minimum *ESS >* 100. In this case, “hidden states” such as the growth rate are estimated alongside static parameters, so also feature in the convergence checks. This means the maximum 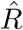 and minimum *ESS* diagnostics are not directly comparable to those of the SIMPLE approach, although they do reflect the computationally expensive aspects of each approach (estimating the static parameters in the SIMPLE approach is much more expensive than estimating the hidden states).

### 2.3 The Abbott approach

The Abbott approach [7] models the first-order difference (other order differences are possible but not considered here) of logit-transformed daily infection incidence using a Laplace eigenfunction approximation [30, 31] to a zero-mean Gaussian process:

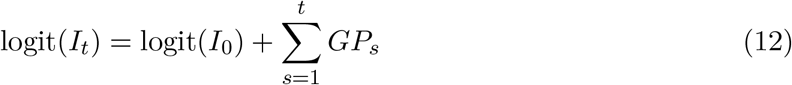

where *I*_0_ is the initial incidence and *GP*_*s*_ is the value of the approximated Gaussian process at time *s*. At small incidence values *I*_*t*_, this is approximately equivalent to modelling the growth rate in incidence 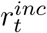 as a Gaussian process:

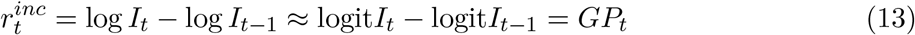

A squared exponential kernel, parameterised by variance 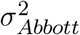 and lengthscale *l*, is used for the Gaussian process:

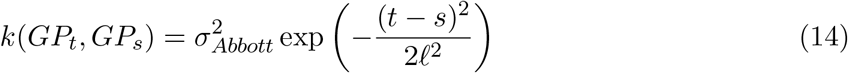

If the lengthscale *l* is large relative to unit time-steps, this Gaussian process can be locally approximated by an AR(1) process: *GP*_*t*+1_ = *ϕGP*_*t*_ + *ϵ*_*t*_ where *ϕ* = exp(−1*/*2*l*^2^) and 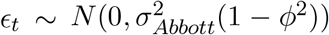. This approximation highlights a mild similarity to our basic model: the model used in the Abbott approach can be (very loosely) viewed as a Gaussian random walk, except on 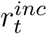 and with a drift towards zero growth. The strength of this drift is controlled by *l* with larger values of *l* indicating a slower drift towards zero growth.

Prevalence *P*_*t*_ is modelled as a convolution of past incidence with a test-sensitivity function that describes how likely an individual is to test positive *u* days after infection (equation 5). During the COVID-19 pandemic, estimates of the test sensitivity function produced by Hellewell, Russell, Matthews, et al. [21] were used. We use the same estimates in our implementation (including in simulated data where applicable), although these curves depend on the pathogen (including variant, in the case of SARS-CoV-2 [32]), testing procedures (e.g. swabbing technique [33] and storage during transport [12]), and the population being tested (e.g. age structure and vaccination status [34]). As a result, these curves are not universal, and should be separately estimated for each scenario. We compare multiple estimates of these curves, and the sensitivity of model results to these curves, in Supplementary Section 5.

The original Abbott approach used a normal likelihood for observed swab positivity with mean *P*_*t*_ and an estimated variance, a necessary simplification given the format of their data. We employ the beta-binomial observation model instead, although the binomial or weighted observation models are also possible and can be implemented by changing only a few lines of the Stan code. The original model also leveraged antibody data, accounting for vaccination status, in their model. We focus on swab positivity data so do not consider this here. Their antibody model could be included in any of the three approaches discussed in this paper. We leave this for future work.

Growth rates are back-calculated from the estimated prevalence by setting *r*_*t*_ = log(*P*_*t*_*/P*_*t−*1_). A weakly informative inverse-gamma prior distribution is used for *l*, a zero-mean normal distribution with standard deviation 0.1 is used as the prior distribution for *σ*_*Abbott*_, a uniform prior distribution is used for *ρ*, a normal prior distribution is used for the initial logit-incidence (at the beginning of the 50-day wind-in period) with mean −4.6 and standard deviation 2, and a zero-mean normal prior distribution with standard deviation 0.25 is used for the initial growth rate in incidence (at the beginning of the 50-day wind-in period).

As the Abbott approach models infection incidence directly, the reproduction number can be estimated by directly applying the renewal model (equation 4) to the estimated incidence curves.

Abbott and Funk provide source code in R and cmdstanr to fit their model. When fitting their model, we use 200 warm-up and 300 sampling iterations. As in the Eales approach, convergence is measured by running three chains and checking that maximum 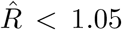 and minimum*ESS >* 100. This source code assumes data of a different format to ours (their inputs are daily means and credible intervals from a different Bayesian model), so we adapt their code to fit to raw data. We also remove code relating to their antibody model. The modified code is provided on GitHub.

### 2.4 Comparison of approaches

Of the three approaches considered in this paper, the SIMPLE approach is mathematically the simplest. The inference method (SMC + PMMH) produces direct samples from the target posterior distribution of a relatively simple model (a random walk). The Eales approach is slightly more complex, relying on splines to smooth between knots, although the inference method (HMC with NUTS) also directly targets the posterior distribution of interest. Finally, the Abbott approach is the most complex, with the inference method (also HMC with NUTS) targeting the posterior distribution of a Laplace eigenfunction approximation of a Gaussian process. For this reason, the SIMPLE and Eales approaches are described as “exact”, while the Abbott approach is “approximate”.

Both the SIMPLE and the Abbott approaches exhibit moderate computational complexity as they both rely on Monte Carlo sampling methods to sample from their target posterior distributions. Despite incorporating methodological improvements and using the same inference method as the Abbott approach, the Eales approach has higher computational complexity due to a greater number of highly-correlated parameters to estimate, whereas the Laplace eigenfunction approximation in the Abbott approach means that sampling from the resulting posterior distribution is substantially easier.

When estimating the growth rate *r*_*t*_, the SIMPLE approach makes relatively few assumptions about the underlying epidemic (that *r*_*t*_ follows a random walk). The Eales approach makes an additional assumption: that the growth rate varies smoothly between knots. The Abbott approach always models incidence (rather than just prevalence), requiring additional assumptions about the generation time distribution and time-varying test sensitivity, thus making more assumptions than the other approaches. When estimating the reproduction number *R*_*t*_, the SIM-PLE approach also makes these additional assumptions. Finally, the Eales approach, with the trailing window of 14 days, makes the most assumptions when estimating *R*_*t*_.

The SIMPLE approach obtains comparable computational efficiency as the Abbott approach while retaining the exactness of the Eales approach. This allows us to fit the SIMPLE models to a wide range of simulated datasets multiple times within a reasonable timeframe. Differences in results between datasets and models can be attributed to real variation, rather than artefacts of mathematical approximations, making it easier to validate the models themselves. A summary of the key differences between the three approaches is provided in Table 1.

**Table 1:** An overview of key differences in the three approaches considered in this paper. Computational complexity is with respect to the precise models considered in this manuscript. Each approach has benefits and drawbacks depending on the specific scenario.

|  | SIMPLE | Eales | Abbott |
| --- | --- | --- | --- |
| Language | Julia | R & Stan (C++) | R & Stan (C++) |
| Inference method | SMC & PMMH | HMC with NUTS | HMC with NUTS |
| Smoothing model | Random walk on $r_t$ or $R_t$ | B-splines on $P_t$ | Gaussian process on $I_t$ |
| Mathematical complexity | Lowest | Moderate | Highest |
| Computational complexity | Moderate | High | Moderate |
| Assumptions made | Fewest ($r_t$) or many ($R_t$) | Few ($r_t$) or most ($R_t$) | Many |
| Exactness | Exact* | Exact* | Approximate |

\*All approaches involve Monte Carlo estimation procedures and thus are subject to Monte Carlo error.

Finally, the hidden-state models used by all three approaches have approximate function space interpretations that reveal underlying assumptions about smoothness. The SIMPLE approach models the growth rate *r*_*t*_ as a first-order random walk, implying a log-prior probability proportional to 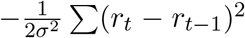, which can be viewed as a discrete approximation of the integral penalty 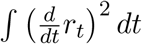, thus implying the equivalent continuous growth rate function lies in the *W* ^1,2^ Sobolev space (the space of functions in *L*^2^ that have square-integrable weak first derivatives). The Eales approach directly penalises the second-order differences of the spline coefficients, which (by the same argument) places logit-prevalence in the *W* ^2,2^ Sobolev space. Since the growth rate is approximately the first derivative of logit-prevalence, this implies that the growth rate function inferred by the Eales approach also lies in the *W* ^1,2^ Sobolev space. Both models, therefore, penalise roughness in a similar fashion.

In contrast, the Abbott approach models the daily change in logit-incidence using a Gaussian process with a squared exponential kernel, the sample paths of which are almost surely infinitely differentiable [35]. Because prevalence is a convolution of incidence, the implied prevalence in this approach inherits the same high degree of smoothness. This is a much stronger assumption than the other two approaches and may be unrealistic for many real-world epidemic processes, which can be subject to abrupt changes due to interventions, the emergence of new variants, and other factors [36]. While outside the scope of this paper, alternative Gaussian process kernels could be used in the Abbott approach (the Matérn kernel is a popular choice with direct links to the function spaces of the SIMPLE and Eales approaches [35]).

### 2.5 Simulated data

Simulated datasets, approximately reflecting the dynamics of COVID-19 in England, are generated by sampling from the prior predictive distribution of a given model with predetermined parameter values. In general, *T* days (default 100) of an epidemic are simulated with a fixed initial prevalence *P*_0_ (default 1%) and growth rate *r*_0_ (default 0). As the Abbott approach models incidence and requires a wind-in period (default 50 days when simulating), we instead fix infection incidence at the start of the wind-in period at 0.1%. Other model-specific parameters are set to default values as outlined in Table 2 unless otherwise stated. These are chosen to reflect estimates from the REACT-1 study, thus representing plausible values for SARS-CoV-2-like epidemics. To prevent unrealistic scenarios, we bound simulated prevalence in the range (0.1%, 30%) by resampling if a simulation exceeds these bounds. Example simulations are shown in Figure 1.

**Table 2:** Default parameter values as listed in this table are used when simulating data, unless otherwise stated. The values are chosen to reflect estimates from fitting each model to the REACT-1 study. Prevalence and incidence are as a proportion of the population.

| Parameter | Default value | Description |
| --- | --- | --- |
| General parameters |  |  |
| $T$ | 100 days | Duration of the simulated epidemic |
| $P_0$ | 0.01 | Initial prevalence |
| $r_0$ | 0 | Initial growth rate (per day) |
| $n_t, t \in \{1, \dots, T\}$ | 5,000 | Daily swabs |
| SIMPLE approach (growth rate epidemic model) |  |  |
| $\sigma$ | 0.01 | Standard deviation of random walk on $r_t$ (per day) |
| $\rho$ | $2 \times 10^{-4}$ | Overdispersion parameter (beta-binomial observation distribution) |
| $\xi$ | 0.5 | Scale of log-normal distribution for simulated weights |
| Eales approach |  |  |
| $\sigma_{Eales}$ | 0.1 | Standard deviation of random walk on spline knots (per knot) |
| $\rho$ | $2 \times 10^{-4}$ | Overdispersion parameter (beta-binomial observation distribution) |
| Abbott approach |  |  |
| $\sigma_{Abbott}$ | 0.08 | Gaussian process kernel amplitude |
| $\ell$ | 10 | Gaussian process kernel lengthscale |
| $\rho$ | $2 \times 10^{-4}$ | Overdispersion parameter (beta-binomial observation distribution) |
| $I_{-init}$ | 0.001 | Initial incidence at the start of the wind-in period |
| $r_{-init}$ | 0 | Initial growth rate in incidence (per day) at the start of the wind-in period |

**Figure 1.**
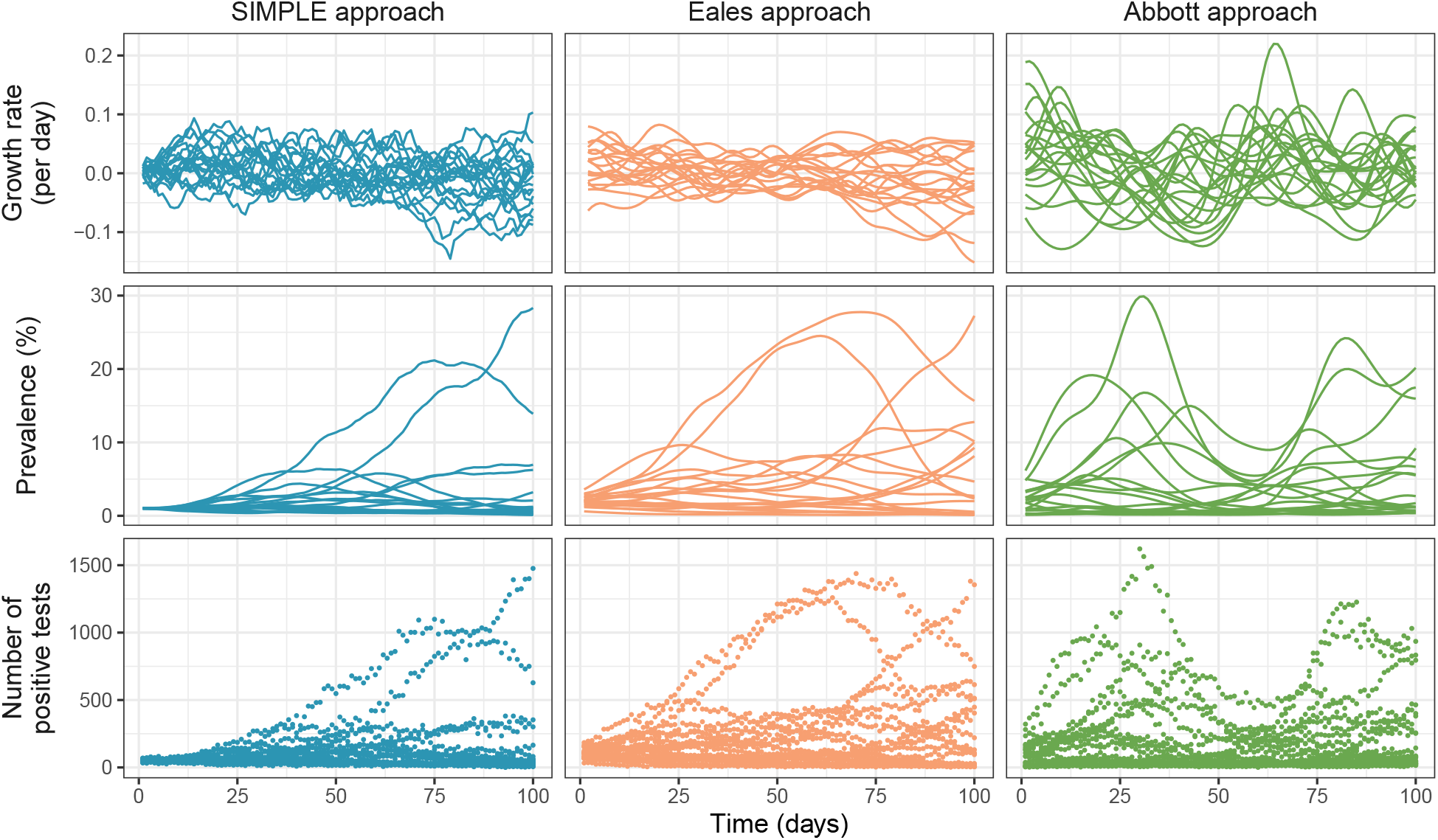
Simulated epidemic trajectories from the SIMPLE, Eales, and Abbott approaches using default parameter values as described in Table 2. A total of 20 simulations are shown per approach, reflecting the range of possible outcomes. The top row shows the simulated growth rate *r*_*t*_, the middle row shows the simulated prevalence *P*_*t*_, and the bottom row shows the simulated number of positive swabs 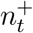.

Weighted survey data are generated by sampling artificial survey weights *w*_*t,i*_ from a log-normal distribution (default log-mean 0 and scale *ξ* = 0.5) and normalising so 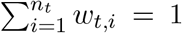. To introduce bias, the probability that individual *i* ∈ {1, …, *n*_*t*_} tests positive is set to:

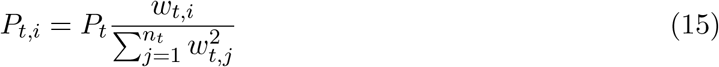

This results in individuals with higher survey weights (i.e., individuals that are underrepresented in the sample) being more likely to test positive, while the weighted average of *P*_*t,i*_ is equal to *P*_*t*_. The simulated dataset then consists of {*x*_*t,i*_, *w*_*t,i*_} where each *x*_*t,i*_ is a realisation of a Bernoulli(*P*_*t,i*_) random variable. This imposes a linear relationship between survey weight and individual swab positivity, so represents a high level of bias for any assumed scale *ξ*. Default *ξ* = 0.5 was chosen to reflect the observed distribution of per-round survey weights in the REACT-1 study. An empirical analysis of these weights is provided in Supplementary Section 6.

To consider the effect of survey design and epidemic dynamics, we fit each of the SIMPLE observation models (basic, extra-binomial, and weighted) to simulated datasets of duration *T* = 100 days from each observation model (with the growth rate epidemic model). Five values of the daily sample size *n*_*t*_ are considered (10, 100, 1,000, 10,000, 100,000) and two values of *σ* (0.008, 0.016). When simulating from the extra-binomial model we assume *ρ* = 2 × 10^*−*4^, and when simulating from the weighted model we assume *ξ* = 0.5. The choice of these values were guided by estimates from the REACT-1 study (see the REACT-1 results for *ρ* and Supplementary Section 6 for *ξ*). Each simulation is repeated 10 times to average over stochasticity. To evaluate model performance, we consider the coverage and average width of the 95% credible intervals for estimated prevalence *P*_*t*_. Equivalent results for the growth rate in swab positivity *r*_*t*_ are presented in Supplementary Section 7.1.

### 2.6 The REACT-1 study

The REACT-1 study was an infection prevalence survey that tested for SARS-CoV-2 infection in England between May 2020 and March 2022 [8]. Conducted over 19 rounds, a total of 2.5 million self-administered throat and nose samples were processed using RT-PCR. Daily swab positivity and sample sizes for all 19 rounds of the REACT-1 study are reported in Figure 2. An average of 6,236 samples were taken on each of the 400 days that sampling occurred, or an average of 3,564 samples per day over the 700 days spanned by the study (from 1 May 2020 to 31 March 2022 inclusive).

**Figure 2.**
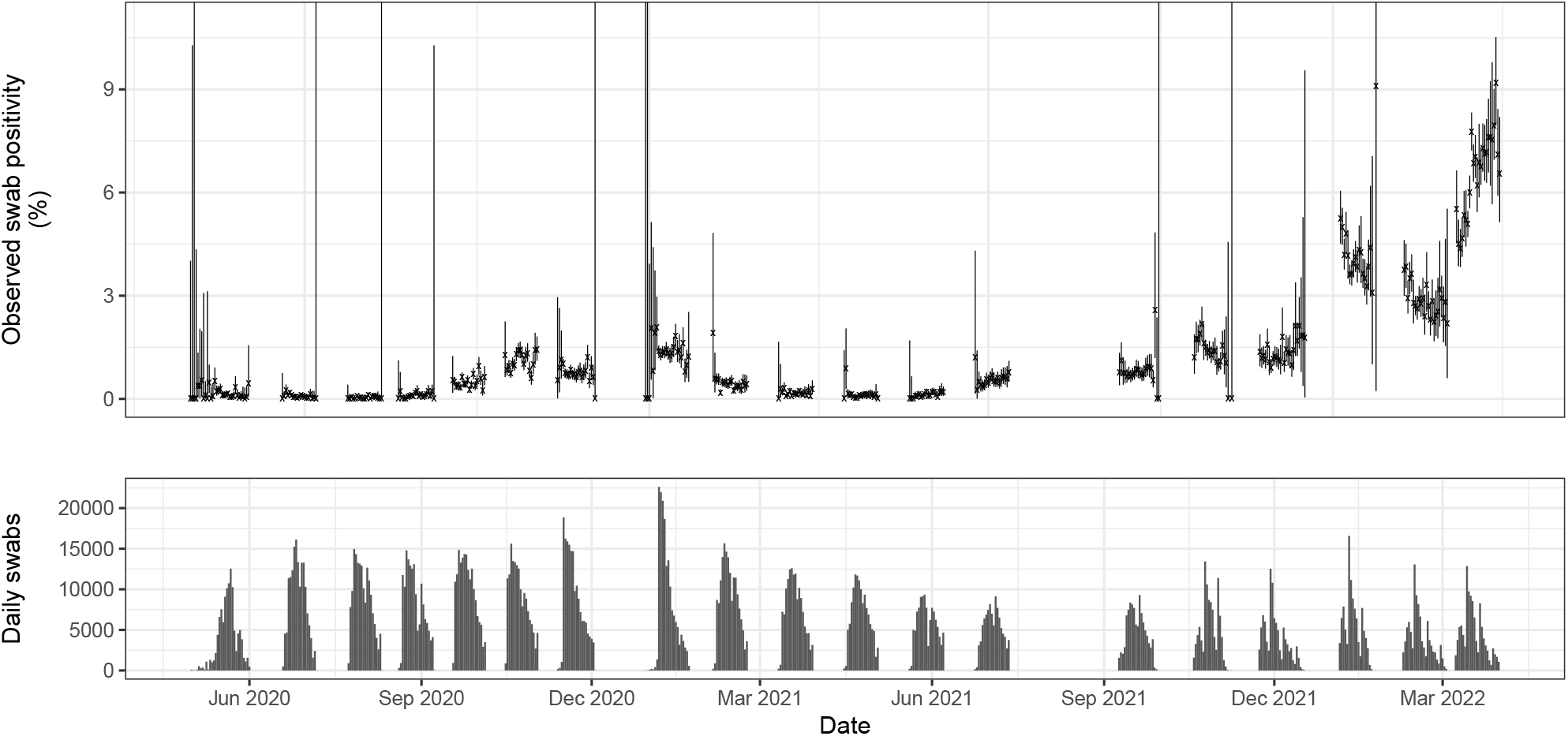
Daily SARS-CoV-2 swab positivity in England from the REACT-1 survey (upper) and corresponding daily sample sizes (lower). Daily 95% confidence intervals (vertical lines) for prevalence were calculated using the Agresti-Coull method [14] in the *binconf* function of the *Hmisc* package in R [37].

## 3 Results

### 3.1 Survey design and epidemic dynamics

In this section we use the SIMPLE approach to investigate the impact of survey design, particularly the number of daily samples and individual response bias, and underlying epidemic dynamics (the variability in the growth rate) on the quality of estimates. Specifically, we consider when each of the three observation models (basic, extra-binomial, and weighted) are appropriate, and how well they perform when fit to simulated data from each model.

All three observation models produce well-calibrated and similarly narrow credible intervals when fit to simulated data from the basic observation model (i.e., a survey with binomial-distributed observations), despite the extra-binomial and weighted observation models featuring an additional (and in this case, unnecessary) parameter (Figure 3, column A). The width of these credible intervals decreases as *n*_*t*_ increases, with 1000 daily samples generally sufficient to produce credible intervals less than 1 percentage point in width, although this is highly simulation-dependent. Larger sample sizes may also allow for the estimation of prevalence by region and/or demographic, as seen in the REACT-1 study. When fit to simulated data with more variable growth rates (higher *σ*, dashed lines), the width of the credible intervals increases slightly, suggesting that a minor increase in daily sample size may be required when the growth rate is changing faster (to achieve the same level of precision). Finally, the weighted observation model produces poor coverage at low sample sizes, due to the breakdown of the normal approximation to the binomial distribution.

**Figure 3.**
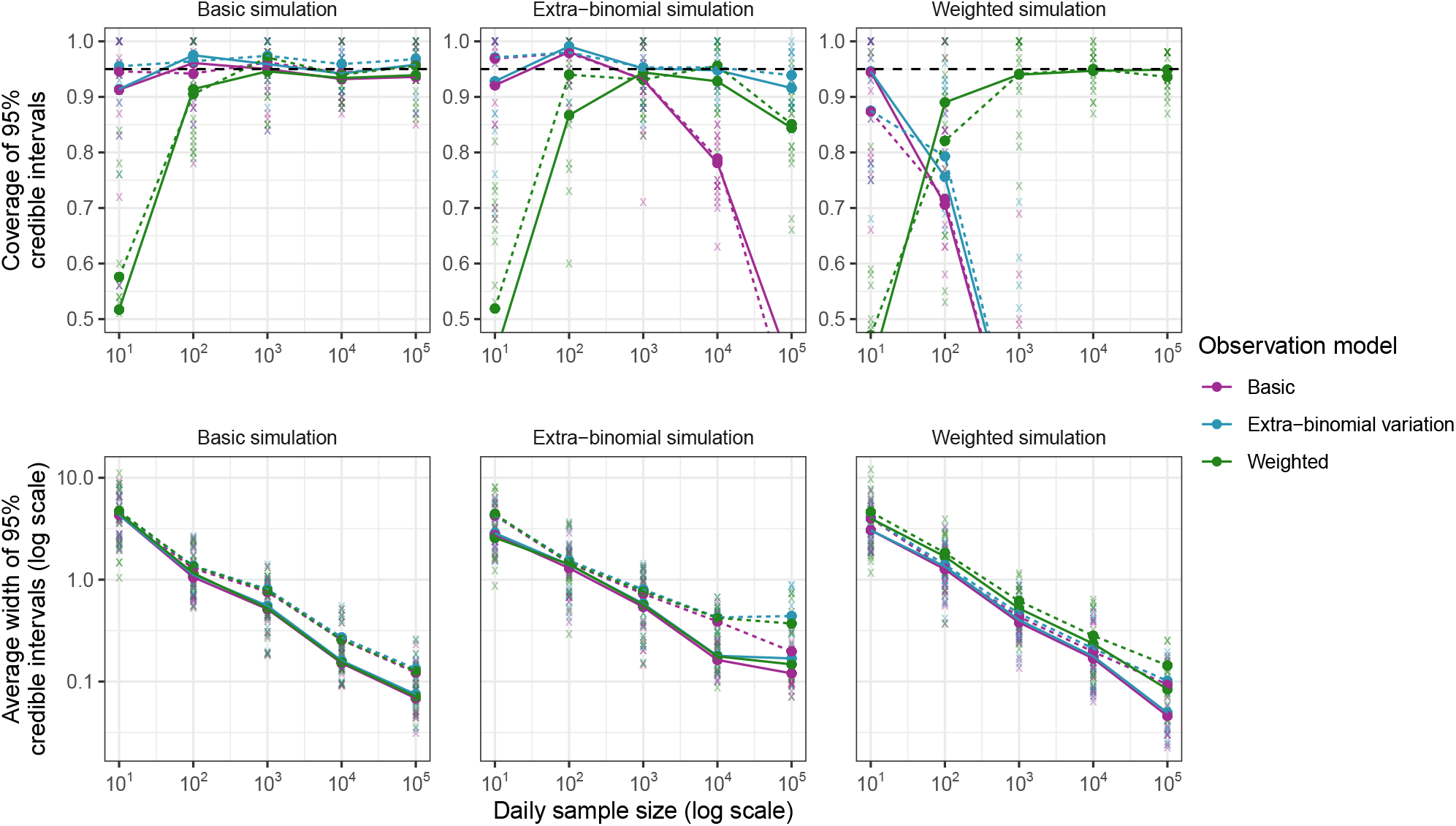
Coverage and average width of 95% credible intervals for prevalence *P*_*t*_ (%) from fitting all three observation models (purple: basic, blue: extra-binomial, green: weighted) to simulated data from each model (column A: basic, column B: extra-binomial, column C: weighted). Results from individual simulations are shown as semi-transparent crosses, with averages over 10 simulations shown as points connected by solid lines (for assumed *σ* = 0.008) and dashed lines (for assumed *σ* = 0.016). A range of assumed daily sample sizes *n*_*t*_ are considered (x-axis). The horizontal black dashed line indicates the target coverage of 0.95. The y-axis for coverage is truncated to (0.5, 1.0), although the coverage in some cases falls outside this range: reaching a minimum average of 0.33 for the basic model fit to the extra-binomial simulations and a minimum average of 0 for the basic and extra-binomial models fit to the weighted simulations (all when *n*_*t*_ = 10^5^).

When fit to simulated data featuring extra-binomial variation, only the extra-binomial model consistently produces well-calibrated estimates of *P*_*t*_ (Figure 3, column B). At larger values of *n*_*t*_, when the normal approximation to the binomial distribution is valid, the weighted model also produces well-calibrated estimates of *P*_*t*_, as the additional parameter *c* allows the model to account for the extra-binomial variation. The basic model, while well calibrated at smaller values of *n*_*t*_, produces poorly calibrated estimates as *n*_*t*_ increases, an example of simple modelling assumptions leading to “model-heavy” inferences. This is due to the basic model assuming that the observation variance is inversely proportional to *n*_*t*_, forcing the credible intervals on *P*_*t*_ to decrease in width as *n*_*t*_ increases, even if additional observation noise is present. The basic model initially accounts for this by artificially increasing the estimated value of *σ*, allowing variation in *r*_*t*_ to capture the additional variation in 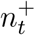 (at the cost of biased estimates of *σ* and *r*_*t*_—see Supplementary Section 7.2), although this breaks down as *n*_*t*_ gets very large. This has real-world implications as seen on the REACT-1 dataset, where the original Eales approach (using a binomial observation model) produces noticeably different estimates of *r*_*t*_ and *P*_*t*_ compared to the modified approach using an extra-binomial observation model (Supplementary Section 4). Finally, both the basic and extra-binomial models perform poorly when fit to simulated weighted data, with only the weighted model being able to recover the true prevalence (Figure 3, column C). Unlike in the other models, estimates of growth rates are less prone to weighting-associated bias (Supplementary Figure S10). We emphasise caution when translating these results to real-world datasets: the simulated weighted model represents an extreme scenario where weights and individual swab positivity are perfectly correlated and the weights are known. In practice, these weights must be estimated through techniques such as random iterative method (RIM) weighting [38], which introduces additional uncertainty that is not accounted for by this model.

### 3.2 Comparing approaches

In this section, we compare the SIMPLE, Eales, and Abbott approaches on 10 simulated datasets from each model with default parameterisations (Table 2), assuming a beta-binomial observation process. All three models produce very similar posterior predictive distributions for swab positivity, and similar posterior distributions for prevalence, suggesting that, given the same observation model, the estimation of *P*_*t*_ and 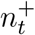 is robust to the differences in smoothing assumptions between models (Figure 4).

**Figure 4.**
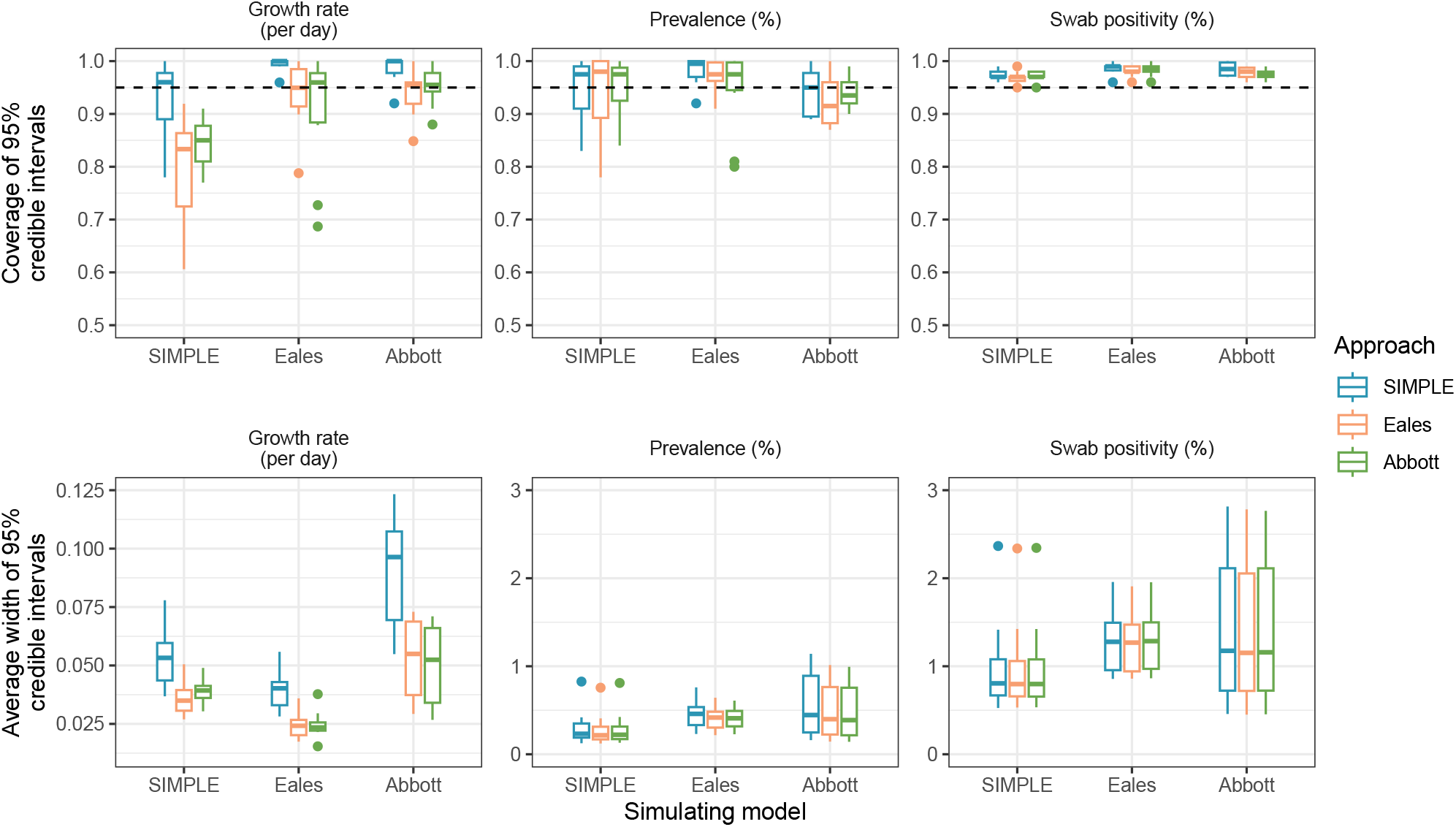
Coverage and average width of 95% credible intervals for the growth rate and prevalence, and coverage and average width of 95% predictive credible intervals for swab positivity. Each approach (SIMPLE, Eales, and Abbott) is fit to 10 simulated datasets from each model. The horizontal black dashed line indicates the target coverage of 0.95. Boxes present the interquartile range of the results with the median shown as a horizontal line. Whiskers extend to the most extreme data point within 1.5 times the interquartile range from the box. Outliers are shown as points.

The models in the Eales and Abbott approaches both enforce a fixed minimum amount of smoothness in the data. In the case of the Eales approach, this is via spline knots being placed less frequently than the observed data; while in the case of the Abbott approach, this results from modelling prevalence as a convolution of smooth incidence and the test-sensitivity function and assuming that the incidence function is infinitely differentiable. This minimum smoothness results in the Eales and Abbott approaches producing narrower credible intervals on *r*_*t*_ than the SIMPLE approach, helpful when the true growth rate is smooth (the SIMPLE approach overcovers *r*_*t*_ in simulations from the Eales and Abbott models), but results in undercoverage when the true growth rate is more variable (Figure 4). These are further examples of simple modelling assumptions leading to “model-heavy” inferences.

The total time taken to fit the models to all 30 simulations was 18m 40s for the SIMPLE (extra-binomial) approach, 54m 58s for the Abbott approach, and 47m 44s for the Eales approach. A successful iteration of the Abbott approach takes less time than the Eales approach, however, the Abbott approach sometimes fails to converge, requiring refitting of the model. Further refinement of the code and/or better selection of prior distributions may improve convergence times for the Abbott approach. A more comprehensive comparison of the runtime of each approach is provided in Supplementary Section 8.

### 3.3 The REACT-1 study

In this section we compare all three approaches on the REACT-1 dataset. First we focus on estimating the growth rate *r*_*t*_ and prevalence *P*_*t*_, and then consider estimation of the reproduction number *R*_*t*_. Assuming fixed values of the static parameters over the entire study period of 700 days may not be appropriate [39]. To assess temporal variation in these parameters, we fit the models separately to four time periods: 1 May 2020 to 3 December 2020 (REACT-1 study rounds 1-to-7), 30 December 2020 to 12 July 2021 (REACT-1 study rounds 8-to-13), 9 September 2021 to 17 December 2021 (REACT-1 study rounds 14-to-16), and 5 January 2022 to 31 March 2022 (REACT-1 study rounds 17-to-19). These periods align approximately with changes in the dominant SARS-CoV-2 variant in England (Wildtype, Alpha, Delta, and Omicron) and large gaps in sampling.

All approaches (when using beta-binomial observation distributions) produce similar estimates of the growth rate *r*_*t*_, prevalence *P*_*t*_, and predictive swab positivity 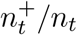, when fit to the REACT-1 dataset (Figure 5), with an average width of 95% credible intervals for *P*_*t*_ of 0.26 to 0.27 percentage points. Minor differences, largely arising from different prior assumptions about the initial growth rate *r*_0_, are observed in estimates at *t* near zero. All approaches also produce very similar estimates of the overdispersion parameter *ρ*, with posterior mean values of 1.9×10^*−*4^ to 2.0 × 10^*−*4^ (Table 3). Other parameters are not directly comparable. Finally, all approaches produced posterior predictive distributions for 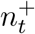 that slightly overcovered the observed data (Table 3). These estimates differ from estimates reported by the REACT-1 study due to our use of a beta-binomial observation model, rather than the binomial observation model used in the original study [17] -we compare these for the Eales approach in Supplementary Section 4.

**Table 3:**
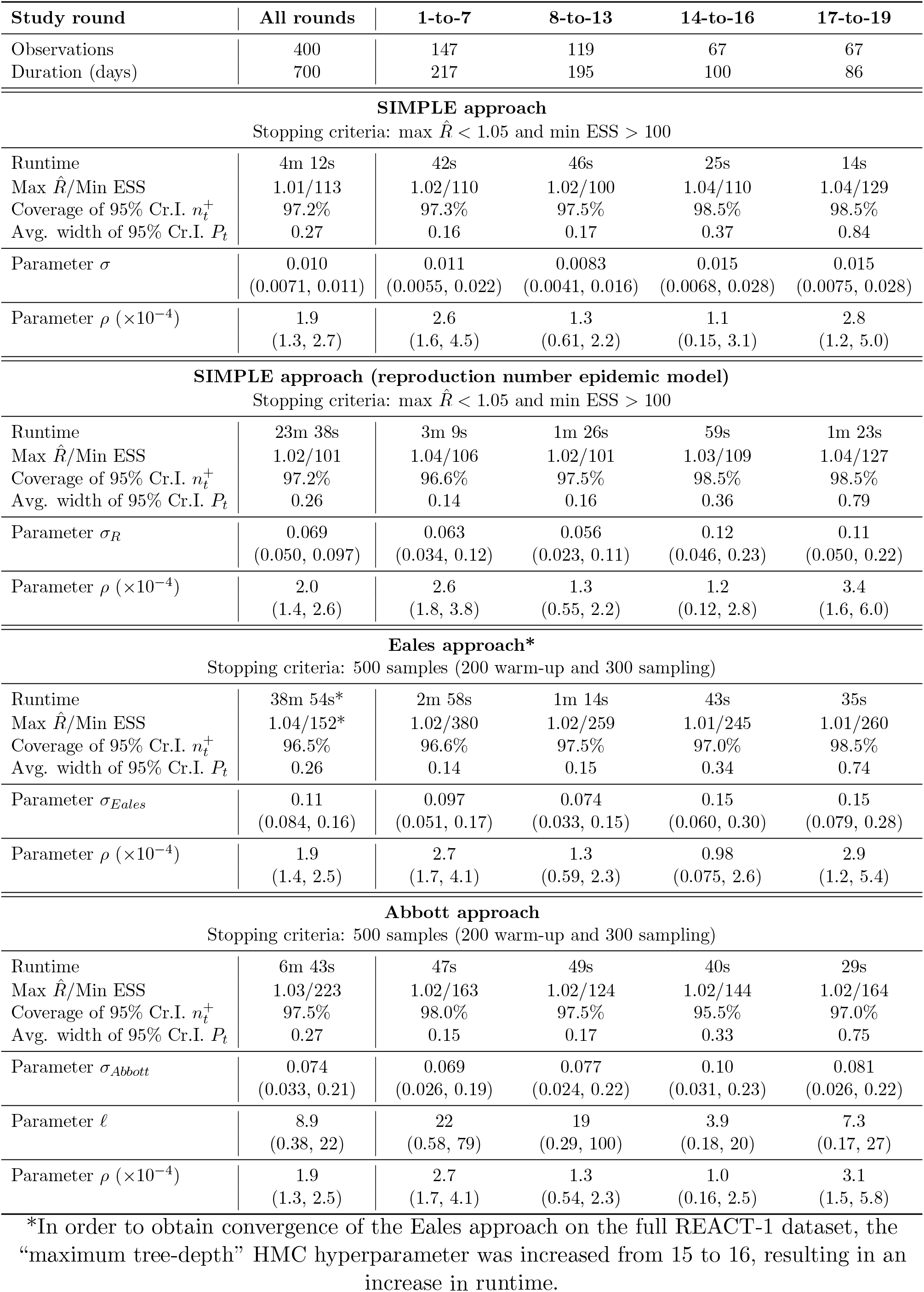
Results from fitting the three approaches to data from the REACT-1 prevalence study. Runtimes were measured once for each dataset considered and can vary considerably. Convergence diagnostics of maximum 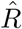 and minimum ESS are reported, although these are not directly comparable between the SIMPLE and Eales/Abbott approaches. Measures of fit are reported as coverage of the posterior predictive distribution and average width of 95% credible intervals on *P*_*t*_ (in terms of percentage points). Parameter estimates are shown as posterior means with 95% credible intervals in parentheses. Note that *σ*_*Eales*_ depends on the knot-spacing, which varies slightly between study rounds, so these estimates are not directly comparable even within the same model.

| Study round | All rounds | 1-to-7 | 8-to-13 | 14-to-16 | 17-to-19 |
| --- | --- | --- | --- | --- | --- |
| Observations | 400 | 147 | 119 | 67 | 67 |
| Duration (days) | 700 | 217 | 195 | 100 | 86 |
| **SIMPLE approach** |  |  |  |  |  |
| Stopping criteria: max $\hat{R} < 1.05$ and min ESS $> 100$ | | | | | |
| Runtime | 4m 12s | 42s | 46s | 25s | 14s |
| Max $\hat{R}$/Min ESS | 1.01/113 | 1.02/110 | 1.02/100 | 1.04/110 | 1.04/129 |
| Coverage of 95% Cr.I. $n_t^+$ | 97.2% | 97.3% | 97.5% | 98.5% | 98.5% |
| Avg. width of 95% Cr.I. $P_t$ | 0.27 | 0.16 | 0.17 | 0.37 | 0.84 |
| Parameter $\sigma$ | 0.010<br>(0.0071, 0.011) | 0.011<br>(0.0055, 0.022) | 0.0083<br>(0.0041, 0.016) | 0.015<br>(0.0068, 0.028) | 0.015<br>(0.0075, 0.028) |
| Parameter $\rho (\times 10^{-4})$ | 1.9<br>(1.3, 2.7) | 2.6<br>(1.6, 4.5) | 1.3<br>(0.61, 2.2) | 1.1<br>(0.15, 3.1) | 2.8<br>(1.2, 5.0) |
| **SIMPLE approach (reproduction number epidemic model)** |  |  |  |  |  |
| Stopping criteria: max $\hat{R} < 1.05$ and min ESS $> 100$ | | | | | |
| Runtime | 23m 38s | 3m 9s | 1m 26s | 59s | 1m 23s |
| Max $\hat{R}$/Min ESS | 1.02/101 | 1.04/106 | 1.02/101 | 1.03/109 | 1.04/127 |
| Coverage of 95% Cr.I. $n_t^+$ | 97.2% | 96.6% | 97.5% | 98.5% | 98.5% |
| Avg. width of 95% Cr.I. $P_t$ | 0.26 | 0.14 | 0.16 | 0.36 | 0.79 |
| Parameter $\sigma_R$ | 0.069<br>(0.050, 0.097) | 0.063<br>(0.034, 0.12) | 0.056<br>(0.023, 0.11) | 0.12<br>(0.046, 0.23) | 0.11<br>(0.050, 0.22) |
| Parameter $\rho (\times 10^{-4})$ | 2.0<br>(1.4, 2.6) | 2.6<br>(1.8, 3.8) | 1.3<br>(0.55, 2.2) | 1.2<br>(0.12, 2.8) | 3.4<br>(1.6, 6.0) |
| **Eales approach\*** |  |  |  |  |  |
| Stopping criteria: 500 samples (200 warm-up and 300 sampling) |  |  |  |  |  |
| Runtime | 38m 54s* | 2m 58s | 1m 14s | 43s | 35s |
| Max $\hat{R}$/Min ESS | 1.04/152* | 1.02/380 | 1.02/259 | 1.01/245 | 1.01/260 |
| Coverage of 95% Cr.I. $n_t^+$ | 96.5% | 96.6% | 97.5% | 97.0% | 98.5% |
| Avg. width of 95% Cr.I. $P_t$ | 0.26 | 0.14 | 0.15 | 0.34 | 0.74 |
| Parameter $\sigma_{Eales}$ | 0.11<br>(0.084, 0.16) | 0.097<br>(0.051, 0.17) | 0.074<br>(0.033, 0.15) | 0.15<br>(0.060, 0.30) | 0.15<br>(0.079, 0.28) |
| Parameter $\rho (\times 10^{-4})$ | 1.9<br>(1.4, 2.5) | 2.7<br>(1.7, 4.1) | 1.3<br>(0.59, 2.3) | 0.98<br>(0.075, 2.6) | 2.9<br>(1.2, 5.4) |
| **Abbott approach** |  |  |  |  |  |
| Stopping criteria: 500 samples (200 warm-up and 300 sampling) |  |  |  |  |  |
| Runtime | 6m 43s | 47s | 49s | 40s | 29s |
| Max $\hat{R}$/Min ESS | 1.03/223 | 1.02/163 | 1.02/124 | 1.02/144 | 1.02/164 |
| Coverage of 95% Cr.I. $n_t^+$ | 97.5% | 98.0% | 97.5% | 95.5% | 97.0% |
| Avg. width of 95% Cr.I. $P_t$ | 0.27 | 0.15 | 0.17 | 0.33 | 0.75 |
| Parameter $\sigma_{Abbott}$ | 0.074<br>(0.033, 0.21) | 0.069<br>(0.026, 0.19) | 0.077<br>(0.024, 0.22) | 0.10<br>(0.031, 0.23) | 0.081<br>(0.026, 0.22) |
| Parameter $\ell$ | 8.9<br>(0.38, 22) | 22<br>(0.58, 79) | 19<br>(0.29, 100) | 3.9<br>(0.18, 20) | 7.3<br>(0.17, 27) |
| Parameter $\rho (\times 10^{-4})$ | 1.9<br>(1.3, 2.5) | 2.7<br>(1.7, 4.1) | 1.3<br>(0.54, 2.3) | 1.0<br>(0.16, 2.5) | 3.1<br>(1.5, 5.8) |

\*In order to obtain convergence of the Eales approach on the full REACT-1 dataset, the “maximum tree-depth” HMC hyperparameter was increased from 15 to 16, resulting in an increase in runtime.

**Figure 5.**
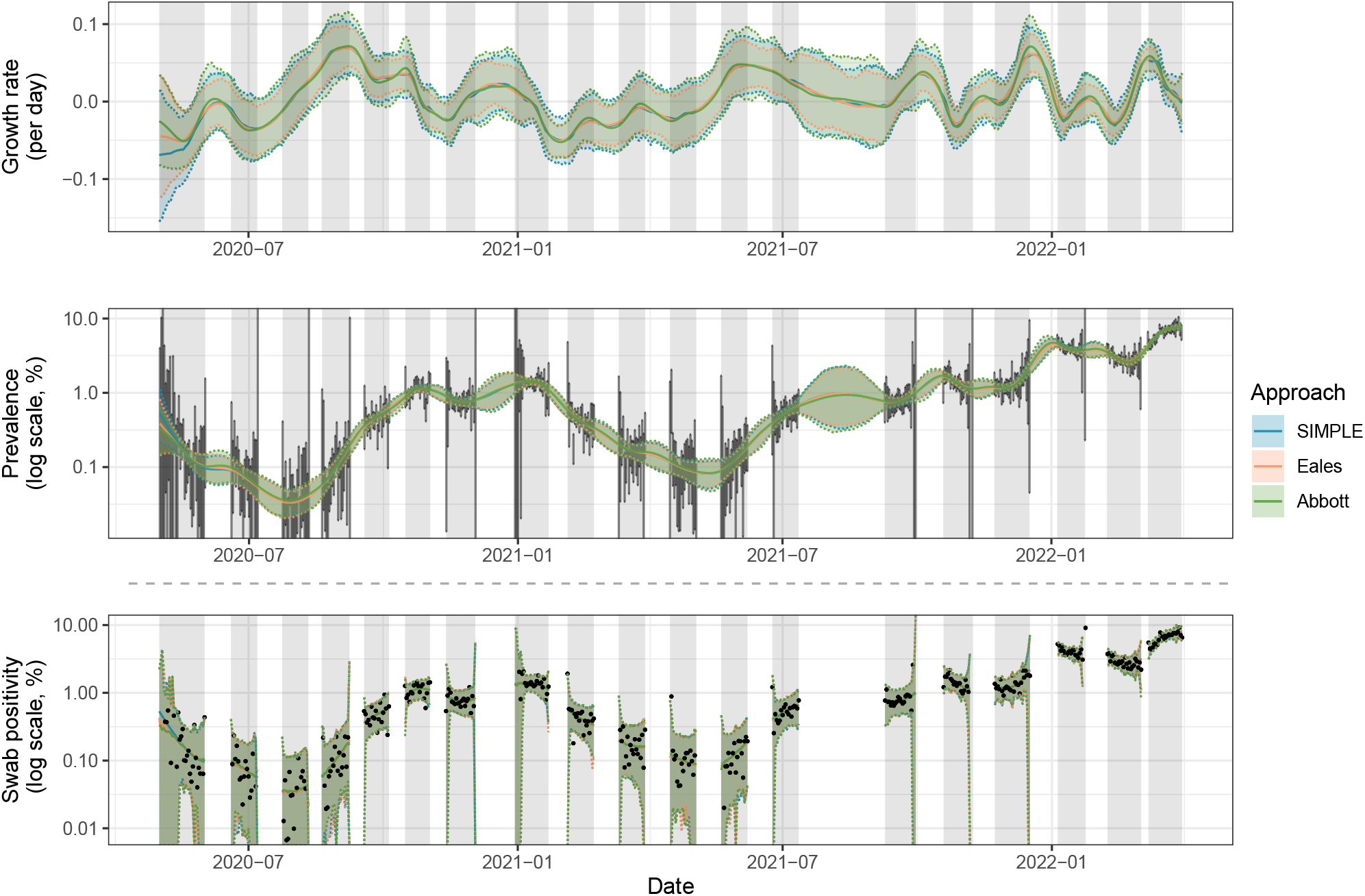
Estimates of the growth rate *r*_*t*_, prevalence *P*_*t*_, and predictive swab positivity 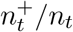, for SARS-CoV-2 in England between 1 May 2020 and 31 March 2022 using data from the REACT-1 study. All three approaches are fit assuming a beta-binomial observation distribution. Solid coloured lines show the posterior means while shading and dashed lines show 95% credible intervals (of the posterior distribution for *r*_*t*_ and *P*_*t*_, and of the posterior predictive distribution for 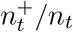). Independent daily confidence intervals from the Agresti-Coull method [14] for *P*_*t*_ are shown in vertical grey lines. The data, daily observed swab positivity 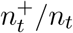, are shown in black points. Grey shading indicates the periods in which sampling was conducted. The predictive distribution for swab positivity depends on the number of swabs taken each day *n*_*t*_, which tends to be lower in the early and late periods of each sampling round, hence the wider credible intervals at the boundaries of each study round.

Results from the SIMPLE approach (with the growth rate epidemic model) suggest that *σ* was higher in study rounds 14-to-16 and 17-to-19 (central estimates of *σ* = 0.015) than in study rounds 1-to-7 (central estimate of *σ* = 0.011) and rounds 8-to-13 (central estimate of *σ* = 0.0083), indicating greater variability in the growth rate in later study rounds. The results also suggest that *ρ* was higher in study rounds 1-to-7 and 17-to-19 (central estimates of 2.6 and 2.8, respectively) than in study rounds 8-to-13 and 14-to-16 (central estimates of 1.3 and 1.1), suggesting greater observation noise at the study’s start and end. These trends are consistent with results from the SIMPLE approach with the reproduction number epidemic model, and the Eales and Abbott approaches (Table 3). While differences in parameter estimates are not statistically significant, estimates of the growth rates *r*_*t*_ do show some sensitivity to whether the models are fit separately to each time period or all together (Supplementary Section 9.1).

The SIMPLE approach (with the growth rate epidemic model) and Abbott approach exhibit comparable runtimes, taking 4m 12s and 6m 43s on the full dataset (all study rounds), or 14s and 29s on the smallest dataset (study rounds 17-to-19), respectively. This is verified in Supplementary Section 8, where a more extensive comparison of runtimes is provided. The Abbott approach can sometimes fail to converge, requiring refitting of the model, thus increasing the total time taken to fit the model. The Eales approach is slower than both of these approaches, particularly on larger datasets, taking 38m 54s to fit to the full dataset and 35s to fit to the smallest dataset. This slowdown is partially due to the need to increase the maximum tree-depth HMC hyperparameter from 15 to 16 for convergence on larger datasets. Finally, the SIMPLE approach with the reproduction number epidemic model is the slowest on smaller datasets, but remains faster than the Eales approach on larger ones (taking 23m 8s on the full dataset and 1m 23s on the smallest dataset). The runtimes reported in Table 3 are for a single run of each approach. A more extensive analysis of runtimes is included in Supplementary Section 8.

Despite each approach producing similar estimates of *r*_*t*_ and *P*_*t*_ and a similar predictive distribution for 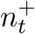 (Figure 5), a number of arbitrary decisions were made when fitting these models. For example, we present estimates of *r*_*t*_ and *P*_*t*_ from fitting to all 19 study rounds simultaneously with constant static parameters, rather than grouping the data into shorter time periods. We also assume the extra-binomial observation model is appropriate, and we do not consider survey weights (as daily-applicable weights were not available). We test these specific modelling decisions in Supplementary Section 9.1, 9.2, and 9.3 respectively. Furthermore, we find evidence for variant-specific parameter values in Supplementary Section 10. Finally, we compare estimates from all approaches with official consensus estimates of the growth rate of COVID-19 in England produced by the UK Health Security Agency (UKHSA) [40] in Supplementary Section 11.

#### The reproduction number

While all approaches produce very similar estimates of *P*_*t*_, and similar estimates of *r*_*t*_ when fit to the REACT-1 data, estimates of *R*_*t*_ differ more substantially (Figure 6). Additional assumptions are required when estimating *R*_*t*_ and this is a quantity known to be sensitive to such assumptions [41]. All models in this section are fit using the extra-binomial observation model.

**Figure 6.**
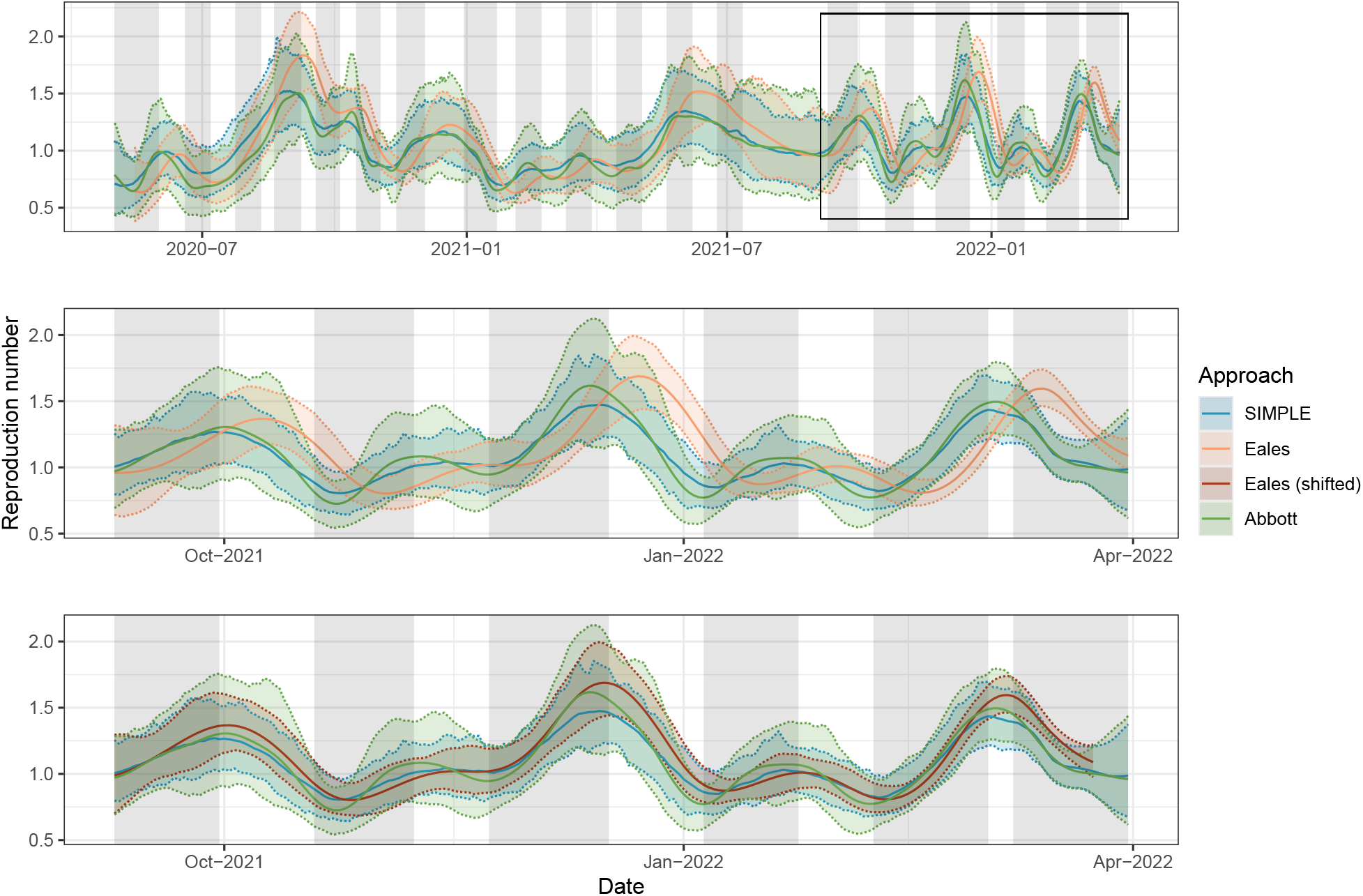
Estimates of the instantaneous reproduction number *R*_*t*_ for SARS-CoV-2 in England between 1 May 2020 and 31 March 2022 using data from the REACT-1 study. All approaches are fit assuming a beta-binomial observation distribution. Solid coloured lines show central estimates while shading and dashed lines show 95% credible intervals. Grey shading indicates the periods in which sampling was conducted. The second panel shows the same estimates for a shorter period (9 September 2021 to 31 March 2022), emphasising the differences between the three approaches. The third panel shows the same estimates, with estimates from the Eales approach shifted by *τ/*2 = 7 days to partially account for bias induced by the trailing window smoothing assumption.

The SIMPLE and Abbott approaches produce similar central estimates of *R*_*t*_, with the Abbott approach producing wider credible intervals on average, reflecting the incorporation of uncertainty in the test-sensitivity function (where the SIMPLE approach uses the mean of the test-sensitivity function). Adapting the SIMPLE approach to allow for uncertainty in this function is possible: by treating the test-sensitivity function as a parameter within PMMH, individual PCR positivity curves can be sampled from estimates in the literature and incorporated into the model. By storing accepted samples of the test-sensitivity function, it is possible to integrate out uncertainty about this function alongside other parameters. We leave the implementation of this to future work.

The Eales approach produces estimates of *R*_*t*_ that are delayed and oversmoothed in comparison to the SIMPLE and Abbott approaches (most apparent in Figure 6-B), a direct result of the trailing window approach to estimating *R*_*t*_. The delay can be partially accounted for by shifting estimates by *τ/*2 days [27] (Figure 6-C), but the oversmoothing is inherent to the approach. Furthermore, as a trailing window of length *τ* = 14 days does not include the entire generation time distribution or test-sensitivity duration, additional biases are introduced. Note that, while the Eales approach was used to estimate *R*_*t*_ in the published results from the REACT-1 study, our estimates may not align with these due to our use of a beta-binomial observation model instead of the original binomial model.

Despite these differences, there is no clear best approach. Most notably, estimates from the SIMPLE and Abbott approaches depend on the assumed test-sensitivity function, which can vary significantly between settings. This is explored further in Supplementary Section 5, where we compare three different curves estimated for SARS-CoV-2 RT-PCR tests, and demonstrate how the choice of curve impacts estimates of *R*_*t*_. We also compare estimates of *R*_*t*_ from these approaches with official consensus estimates produced by the UKHSA [40] in Supplementary Section 11.

As the reproduction number epidemic model in the SIMPLE approach is no longer Markovian, we must use fixed-lag resampling during the parameter inference stage [16], leading to an increase in computation time (23m 38s for the full dataset or 1m 23s on the smallest dataset). The faster runtime of the SIMPLE approach with the growth rate epidemic model could be obtained by fitting a Markovian model (instead of the renewal model), such as a compartmental susceptible-infectious-recovered-type model, which has the added benefit of not requiring a test-sensitivity function. However, this type of model places additional assumptions on the underlying epidemic dynamics, which can be difficult to validate. We leave this for future work.

## 4 Discussion

Smoothing prevalence data allows multiple days to inform point estimates, reducing noise and increasing the confidence in estimates. However, all smoothing methods make assumptions, whether explicitly or implicitly, about the underlying process, and results can be sensitive to these assumptions.

A key concern highlighted in this paper is the presence of overdispersion in the observed data. As shown in Section 3.1, not properly accounting for this can lead to biased prevalence estimates and credible intervals with poor coverage. Growth rate estimates also become more variable, as the model attempts to explain extra-binomial noise through artificial variability in *r*_*t*_. In addition to the simulated results presented in this paper, we also find that this impacts real-world estimates of *P*_*t*_ and *r*_*t*_ in the REACT-1 study. An explicit comparison is given in Supplementary Section 4, emphasising that results can vary considerably, particularly in earlier study rounds. Once overdispersion is appropriately handled, all approaches produce accurate prevalence estimates, even when fit to data generated by other models. This is because prevalence estimates are largely driven by the observed data, not by the smoothing assumptions.

Even when the data are not generated by a beta-binomial sampling process, the overdispersion parameter *ρ* serves as a general error term that can capture unmodelled heterogeneities and sources of variance. The similarity of estimates of *ρ* between the approaches (when fit to real-world data from the REACT-1 study) suggests that this parameter is estimable from observed data independently of the assumed smoothing mechanism. Growth rate estimates (and to a lesser extent, prevalence estimates) in the REACT-1 study were sensitive to the inclusion of this parameter. As the allowance for overdispersion, even when simulated data were generated from a binomial model, had little impact on the accuracy of estimates, we recommend including this parameter in all models. If survey weights are available and the daily sample size is sufficiently large (typically *n*_*t*_ *>* 100, although low-prevalence scenarios may require more), then the weighted model is able to account for both overdispersion and sampling bias, and thus should be the first choice. Unfortunately, without access to survey weights applicable on a daily basis, we were unable to apply this model to the REACT-1 study.

In addition to smoothing prevalence data, the approaches presented in this paper allow for the estimation of the growth rate in prevalence *r*_*t*_. Unlike estimates of prevalence, assumed smoothing dynamics have a substantial impact on estimates of *r*_*t*_. Only the SIMPLE approach produced 95% credible intervals with consistently good coverage of this quantity, at the cost of substantially wider intervals. However, as prevalence is naturally a convolution of past incidence, the additional smoothness implied by the SIMPLE approach with the reproduction number epidemic model, or the Abbott approach, may provide an appropriate way to reduce estimated uncertainty about *r*_*t*_.

Given a predetermined model structure, uncertainty about *r*_*t*_ and *P*_*t*_ arises from two sources: (1) process and observation noise in the epidemic and observation model, and (2) uncertainty about the static parameters governing these models. While uncertainty associated with the former can only practically be reduced by collecting more data, uncertainty about static parameters can be decreased by using more informative prior distributions. At smaller values of *n*_*t*_, a wide uniform prior distribution can result in the posterior distribution assigning probability mass to implausibly large values of *σ* and *ρ*, leading to overestimation of the width of credible intervals for *r*_*t*_ and *P*_*t*_. Using more informative prior distributions that place less prior mass on implausibly large values can help to reduce the width of the credible intervals for *r*_*t*_, *P*_*t*_, and 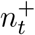, particularly at smaller values of *n*_*t*_. This is demonstrated in Supplementary Section 1.2.

Estimating *R*_*t*_ from prevalence data requires assumptions about the relationship between incidence and swab positivity. This can be achieved by assuming *R*_*t*_ is constant over a sufficiently long time period (Eales approach), by incorporating a test-sensitivity function (SIMPLE and Abbott approaches), or by fitting a fully mechanistic model (not considered here). Each approach introduces different sources of potential bias. The trailing window assumption in the Eales approach guarantees some degree of misspecification [26], which can only be partially mitigated by shifting estimates by *τ/*2 days [27]. Test-sensitivity functions offer an alternative but also vary substantially: several distinct estimates of this function for SARS-CoV-2 RT-PCR tests were produced during the COVID-19 pandemic [21, 33, 42, 43], with meaningful differences in both duration and peak sensitivity. Ideally this function would be estimated for each epidemiological setting being modelled, which would require repeat swabs from a subset of survey participants, although collecting sufficient data may be infeasible in low-prevalence scenarios.

An alternative way to relate incidence to prevalence is to model individual-level cycle threshold (Ct) values [34]. Instead of relying on an assumed test-sensitivity function, this approach fits a model of Ct values as a function of time since infection, replacing the observation model for aggregated data with one for individual Ct data. Models for individual Ct value trajectories have previously been estimated from cross-sectional data, without needing repeated sampling from individuals. This method makes fuller use of the information contained in Ct values, rather than reducing test results to binary outcomes, but requires access to individual-level data, is computationally intensive, and depends on correctly specifying the Ct model.

Our SIMPLE approach is mathematically simpler than the Eales and Abbott approaches, requires no external software (e.g. Stan), is more computationally efficient than the Eales approach, and avoids the approximations used in the Abbott approach. This computational efficiency allows for the rapid testing of a range of models, as demonstrated in Supplementary Section 9.1, where we use the SIMPLE approach to fit a range of models to the REACT-1 dataset. We also demonstrate an adaptation that allows the SIMPLE approach to estimate variant-specific growth rates while leveraging all collected data (including unsequenced samples) in Supplementary Section 10. Finally, the sequential nature of SMC allows for easy modification for online inference, where replacing PMMH with an *SMC*^2^ algorithm [44] would allow for real-time inference as new data are collected without the need to refit the model from scratch.

There are two main limitations to the SIMPLE approach. First, the resampling step in the PMMH algorithm is computationally expensive. For Markovian models, past-state resampling can be disabled during PMMH, enabling fast inference. For non-Markovian models, past-state resampling is necessary, slowing down the PMMH algorithm, as seen in the SIMPLE model for the reproduction number. In these cases, the Abbott approach is more computationally efficient. Second, PMMH relies on stochastic likelihood estimates, whose variance increases with data length and model complexity. This results in *O*(*T* ^2^) time complexity for parameter inference, which was not a bottleneck for the datasets used here (*T* ≤ 700) but will become important for longer time series. While we use relatively simple models in this paper, more complex models would benefit from more advanced algorithms [45].

While we make several improvements to the Eales and Abbott approaches—including using beta-binomial observation distributions and modifying HMC hyperparameters—further optimisation is likely possible which could improve their relative performance. In particular, the Abbott approach sometimes fails to converge. Reparameterising the model, or using better-specified initial values, could reduce or prevent the occurrence of this. In the Abbott approach, we also recommend considering alternative, less smooth, Gaussian process kernels, as the squared exponential kernel is known to produce overly smooth estimates [46]. The very smooth kernel used in the Abbott approach can be partially compensated for by estimating a more variable growth rate *r*_*t*_, which is why simulated data from the Abbott model (Figure 1) exhibits *r*_*t*_ values of greater magnitude than the SIMPLE and Eales approaches, on average.

Estimate precision can be improved by leveraging additional data sources. The original Abbott approach, for example, included an observation model for antibody data [7]. If individual-level data are available, the approach of Pouwels, House, Pritchard, et al. [9], later refined in [47], can be used to model individual probabilities of testing positive. This approach performs post-stratification of individual-level estimates by demographic-geographical response types and has several advantages: multilevel regression and poststratification (MRP) has been shown to out-perform classical survey weighting [48], and partial pooling improves estimation of demographic effects (whereas the approaches presented in this paper must produce demographic-specific estimates by fitting separate models). Future work could explore how to combine the approaches and lessons presented in this paper with MRP-type approaches to improve estimates of *P*_*t*_ and *r*_*t*_.

Our findings indicate that while all three approaches provide robust estimates of prevalence and observed positive swabs, minor differences in their smoothing assumptions can impact growth rate estimates. Applying these approaches to data from the REACT-1 study, we observed that all three produced similar estimates of prevalence and growth rates, with the SIMPLE and Abbott approaches demonstrating greater computational efficiency. By presenting these approaches in common and general notation, alongside well-documented code online, this paper offers a suite of validated tools that can be readily adapted for future epidemic surveys.

## Data availability statement

All source code and data (simulated and from the REACT-1 survey) used in this study are available at: https://github.com/nicsteyn2/EpidemicSurveySmoothing.

