## Supplementary Material for "A Bayesian model for repeated cross-sectional epidemic prevalence survey data"

**3 Eales approach knot-spacing and equivalence with the SIMPLE approach 8**

**4 Comparisons with the original Eales approach 10**

**5 RT-PCR test sensitivity curves for SARS-CoV-2 13**

**6 REACT-1 survey weights 17**

**10 Accounting for multiple variants 27**

**11 Comparisons with UKHSA consensus estimates 30**

### 1 Sensitivity of the SIMPLE approach to choice of prior distributions

#### 1.1 Invariance to reasonable prior distributions

In the main paper, we claim that results from the SIMPLE approach are largely invariant to the reasonable choice of initial distribution for the growth rate $r_0$, initial distribution for prevalence $P_0$, and prior distribution for $\sigma$. We demonstrate this here by refitting the basic model (growth rate epidemic model and basic observation model) to a single realisation of simulated data using different prior distributions. Figure S1 supports our claim. Small deviations in $r_t$ estimates at early time steps are observed when varying the prior for $r_0$, and similarly for $P_t$ when varying the prior for $P_0$. The fixed-lag resampling ensures that future data still influences these estimates, hence they do not vary as much as might be expected. The remaining results are nearly identical regardless of prior distribution. These results hold over repeated simulations.

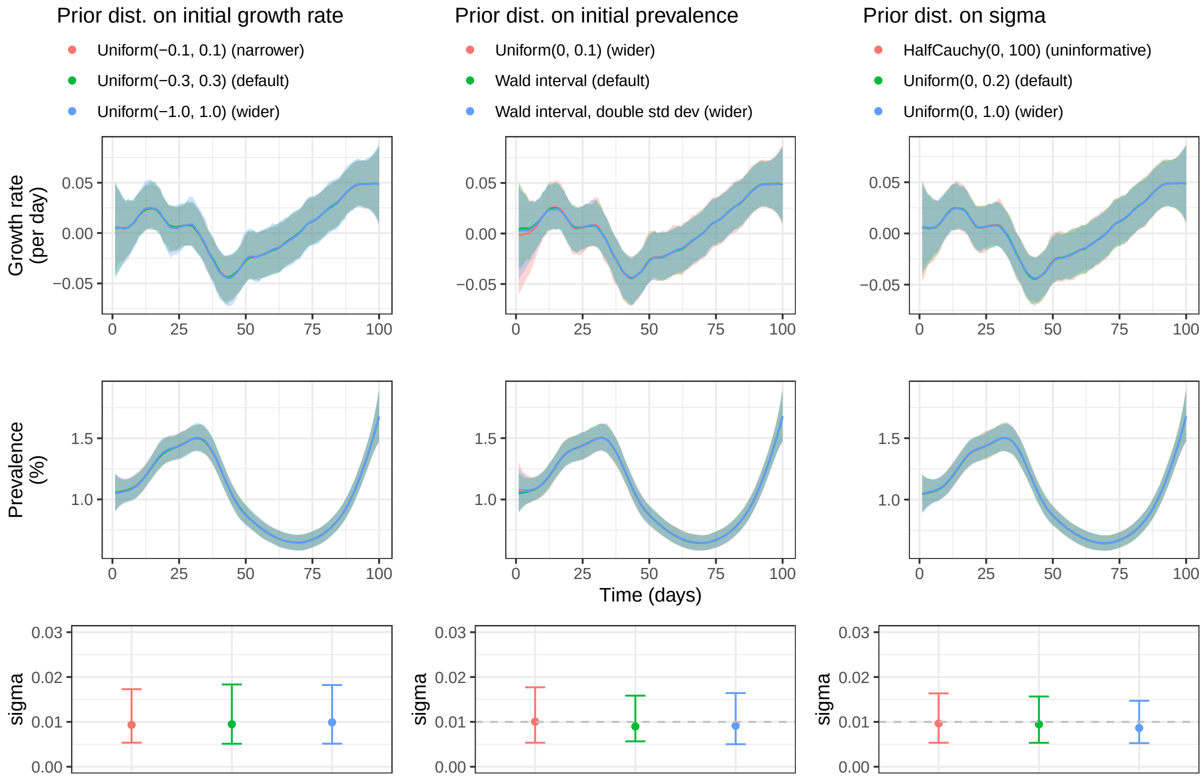

Figure S1: Estimates of the growth rate and prevalence over time (central lines show the posterior mean and shading shows the 95% credible interval), and posterior mean and 95% credible intervals on parameter $\sigma$ when fitting the SIMPLE approach (growth rate epidemic model and binomial observation model) to simulated data. Column 1 shows results under varying prior distributions on the initial growth rate, column 2 shows results under varying prior distributions for the initial prevalence, and column 3 shows results under varying prior distributions for the smoothing parameter $\sigma$.

#### 1.2 Informative prior distributions

In the main paper, we claim that "using more informative prior distributions that place less mass on implausibly large values (of $\sigma$ and $\rho$) can help to reduce the width of the credible intervals" on the hidden states. We demonstrate this here by fitting the beta-binomial model to 10 realisations of simulated data (with $\sigma = 0.008$ and $\rho = 2 \times 10^{-4}$) at various values of $n_t$. First, we fit the model using the default prior distributions ($P(\sigma) \sim U(0, 0.2)$ and $P(\rho) \sim U(0, 0.01)$). Then, we refit the model (to the same simulations) using tighter prior distributions $P(\sigma) \sim U(0.004, 0.016)$ and $P(\rho) \sim U(1 \times 10^{-4}, 4 \times 10^{-4})$, chosen to reflect typical lower and upper 95% credible intervals of the posterior distributions when the default model is fit to simulated data with large $n_t = 10,000$. Finally, we refit all models with $\sigma$ and $\rho$ fixed at their true values (i.e., $P(\sigma) \sim \delta(0.008)$ and $P(\rho) \sim \delta(2 \times 10^{-4})$ where $\delta$ is the Dirac delta function).

Figure S2 presents the coverage and average width of the 95% credible intervals for the growth rate $r_t$ and prevalence $P_t$ (in %) for each of the three scenarios. The effect is most pronounced for growth rate estimates, where credible intervals produced using the tighter prior distribution are substantially narrower than those produced using the default prior distribution, particularly at smaller values of $n_t$ (where there are fewer data to inform the posterior distribution). Using known parameters results in a further reduction in the width of the credible intervals. This effect primarily occurs due to eliminating the possibility of higher values of $\sigma$. The coverage of the credible intervals also improves (becomes closer to 0.95 on average) as removing the possibility of higher values of $\sigma$ also reduces the chance of overcovering the true value (due to wider credible intervals). The effect on prevalence estimates is less pronounced but still noticeable. These improvements are only expected when the prior distributions contain the true values of the parameters.

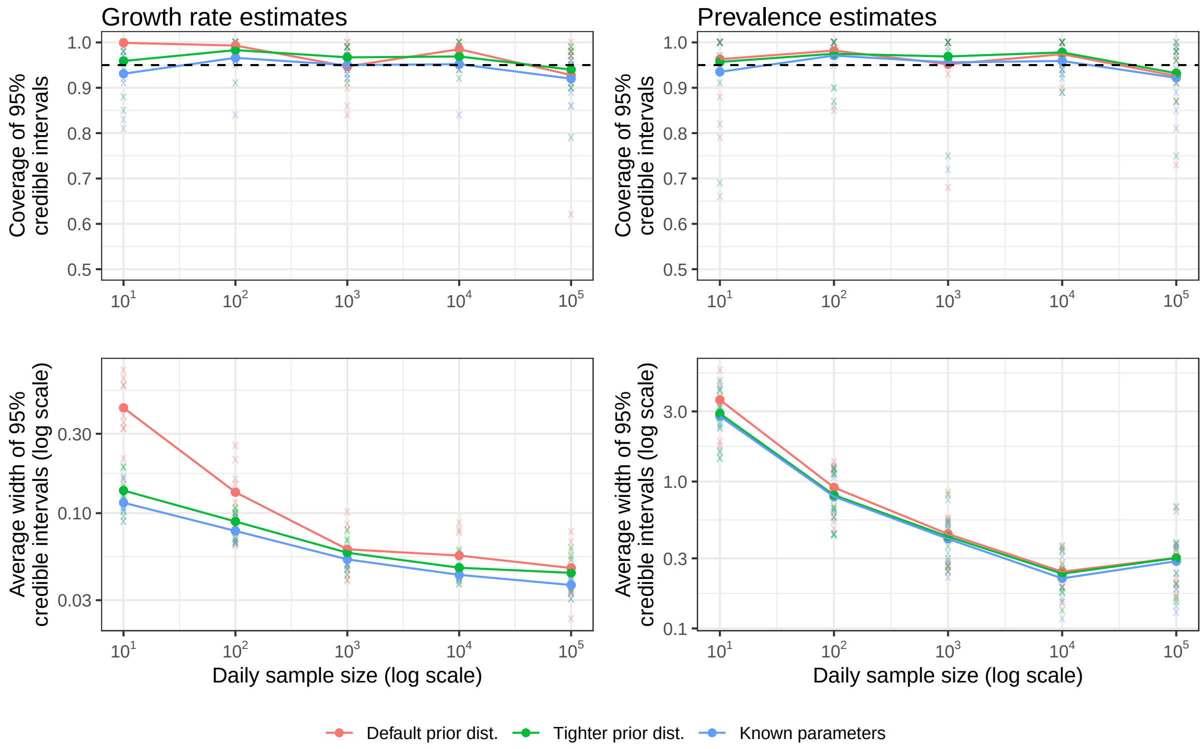

Figure S2: Coverage and average width of 95% credible intervals for the growth rate $r_t$ and prevalence $P_t$ (%) from fitting the beta-binomial model with the default prior distribution (orange), with a tighter prior distribution (green), and with known parameter values (blue). Results from individual simulations are shown as transparent crosses, with averages over 10 simulations shown as points connected by solid lines. A range of assumed daily sample sizes $n_t$ are considered (x-axis). The horizontal dashed line shows the target coverage of 95%.

#### 2 Algorithms

We provide here a concise description of the algorithms used in the SIMPLE approach. We direct readers new to SMC methods to [1], which provides a comprehensive overview of the methodology in an epidemiological context.

For generality, we denote the hidden states at time step $t$ by $X_t$ (e.g. $X_t = (r_t, P_t)$ in the basic model), observed data by $y_t$ (e.g. $y_t = (n_t, n_t^+)$), and model parameters by $\theta$ (e.g. $\theta = (\sigma, \rho)$ in the overdispersed model).

##### 2.1 The bootstrap filter

Given a fixed lag $L$, we approximate the marginal posterior distribution of the hidden-states at time $t$ given past and future observed data $P(X_t|y_{1:t+L}, \theta)$ using a set of $N_x$ "particles" $\{x_t^{(i)}\}_{i=1}^{N_x}$, where each $x_t$ represents a sample from $P(X_t|y_{1:t+L}, \theta)$. The lag $L$ is chosen to be large enough such that $P(X_t|y_{1:t+L}) \approx P(X_t|y_{1:T})$, but small enough to prevent particle degeneracy.

The particles $x_t^{(i)}$ are constructed using a fixed-lag bootstrap filter, described in algorithm 1. Starting with a set of particles sampled from an initial state distribution $\{x_0^{(i)}\}_{i=1}^{N_x} \sim P(X_0)$, we iteratively propagate these particles by first sampling proposed particles from the state-space transition distribution $\{\tilde{x}_t^{(i)}\}_{i=1}^{N_x} \sim P(X_t|x_{t-1}^{(i)}, \theta)$, weighting them according to the observation distribution $w_t^{(i)} = P(y_t|\tilde{x}_t^{(i)}, \theta)$, and then resampling the proposed particles with replacement according to these weights. This process is repeated for each $t$ until the final time step $T$ is reached. The resampling step also resamples particles in the preceding $L$ time steps, ensuring that the particles represent samples from the "smoothing posterior" $\{x_t^{(i)}\}_{i=1}^{N_x} \sim P(X_t|y_{1:t+L}, \theta)$.

In addition to producing the smoothing distribution (conditional on parameters $\theta$), the bootstrap filter also produces estimates of the model likelihood $P(\theta|y_{1:T})$ as a by-product. Specifically, note that the likelihood of the one-step-ahead-forecast can be written as, and approximated by:

$$P(y_t|y_{1:t-1}, \theta) = \int P(y_t|X_t, \theta)P(X_t|y_{1:t-1}, \theta)dX_t \approx \frac{1}{N_x}\sum_{i=1}^{N_x} w_t^{(i)}$$

The model log-likelihood can therefore be written as and then approximated by:

$$\ell(\theta|y_{1:t}) = \sum_{t=1}^{T} \log P(y_t|y_{1:t-1}, \theta) \approx \sum_{t=1}^{T} \log\left(\frac{1}{N_x}\sum_{i=1}^{N_x} w_t^{(i)}\right) \tag{S1}$$

The quality of the approximation improves with larger $N_x$.

**Algorithm 1** Fixed-lag bootstrap filter

1. Input:
   - $N_x$ - number of particles
   - $L$ - fixed lag
   - $\theta$ - parameter vector
   - $P(X_0)$ - initial state distribution
   - $P(X_t|X_{1:t-1}, \theta)$ - state-space transition distribution
   - $P(y_t|X_t, \theta)$ - observation distribution
2. Initialise:
   (a) For $i = 1, ..., N$ sample $x_0^{(i)} \sim P(X_0)$
   (b) Set $t = 1$
3. Project and filter:
   (a) For $i = 1, ..., N$ sample $\tilde{x}_t^{(i)} \sim P(X_t|x_{t-1}^{(i)}, \theta, \cdots)$.
   (b) Calculate weights $w_t^{(i)} = P(y_t|\tilde{x}_t, \theta, \cdots)$
4. Resample:
   (a) Resample $(x_{t-L:t-1}^{(i)}, x_t^{(i)})$ with replacement from $(x_{t-L:t-1}^{(j)}, \tilde{x}_t^{(j)})$ with probability proportional to $w_t^{(j)}$.
5. If $t < T$, increment $t$ and go to step 3. Otherwise, end.

#### 2.2 The particle marginal Metropolis-Hastings algorithm

PMMH is a simple Markov chain Monte Carlo (MCMC) algorithm that allows for the estimation of model parameters $\theta$ given observed data $y_{1:T}$. The algorithm uses the bootstrap filter to estimate the model likelihood, and incorporates these estimates into a Metropolis-Hastings step to determine whether to accept or reject proposed values of $\theta$. We outline this in algorithm 2.

**Algorithm 2** Particle Marginal Metropolis-Hastings

1. Input:
   - A fixed-lag bootstrap filter (and corresponding parameters)
   - $P(\theta)$ - prior distribution for parameters
   - $q(\theta'|\theta)$ - proposal distribution for parameters
   - $ESS$ - target effective sample size
   - $\hat{R}$ - target R-hat statistic
2. Initialise:
   (a) Sample $\theta_0 \sim P(\theta)$
   (b) Run algorithm 1 to obtain estimated log-likelihood $\hat{\ell}(\theta_0|y_{1:T})$
   (c) Set counter $i = 1$
3. Metropolis-Hastings step:
   (a) Sample $\theta' \sim q(\theta'|\theta_{i-1})$
   (b) Run algorithm 1 to obtain estimated log-likelihood $\hat{\ell}(\theta'|y_{1:T})$
   (c) Calculate acceptance probability $\alpha_i$ (equation S2)
   (d) Set $\theta_i = \theta'$ with probability $\alpha_i$, otherwise set $\theta_i = \theta_{i-1}$
4. If $\text{mod}(i, 100) = 0$ AND $ESS_i > ESS$ AND $\hat{R}_i < \hat{R}$, return $\{\theta_i\}$, else GOTO 3.

The acceptance probability $\alpha_i$ at step $i$ is calculated as:

$$\alpha_i = \min\left(1, \frac{\hat{P}(y_{1:T}|\theta')P(\theta')q(\theta_{i-1}|\theta')}{\hat{P}(y_{1:T}|\theta_{i-1})P(\theta_{i-1})q(\theta'|\theta_{i-1})}\right) \tag{S2}$$

where $\theta_{i-1}$ is the previously accepted value of $\theta$, $\theta'$ is the proposed value of $\theta$, and $\hat{P}(y_{1:T}|\theta)$ is the estimated likelihood of the model given the data, calculated using equation S1. $q(\cdot|\cdot)$ denotes the proposal distribution (we use the adaptive multivariate normal approach from [1]) and $P(\theta)$ denotes the prior distribution for $\theta$.

When using the fixed-lag bootstrap filter to estimate the model likelihood, we typically use $N_x = 1000$ particles, chosen to keep the standard deviation of log-likelihood estimates below approximately 1.2 [2]. When fitting to longer time-series, such as the full REACT-1 dataset, we increase this to $N_x = 2000$. The algorithm is halted when the effective sample size (ESS) exceeds 100 and the Gelman-Rubin statistic ($\hat{R}$) is below 1.05. To reduce computation time, these criteria are checked every 100 iterations. We typically run three chains in parallel.

##### 2.3 Marginal posterior distributions

Algorithm 1 produces samples from the conditional posterior distribution $P(X_t|y_{1:t+L}, \theta)$ while algorithm 2 produces samples from the marginal posterior distribution for the model parameters $P(\theta|y_{1:T})$. Particles representing the marginal posterior distribution $P(X_t|y_{1:t+L})$ are obtained by repeatedly sampling $\theta^* \sim P(\theta|y_{1:T})$ and then running algorithm 1 with this value of $\theta^*$. This process is repeated, and the resulting particles are combined to approximate the marginal posterior distribution $P(X_t|y_{1:T})$. By default, we use 100 samples of $\theta$ and $N_x = 2000$ particles per filter, resulting in $2 \times 10^5$ particles representing the marginal posterior distribution.

The choice of fixed-lag $L$ also depends on the application. When fitting a Markovian epidemic model (such as the model for the growth rate $r_t$), we can disable fixed-lag resampling while estimating the model likelihood to speed up computation. When estimating the marginal posterior distribution we use $L = 50$ by default, increasing to $L = 70$ when fitting to the full REACT-1 dataset to allow the model to "bridge the gap" between the end of study round 13 and the start of study round 14. When fitting a non-Markovian epidemic model (such as the model for $R_t$) we must choose $L$ to include the majority of the mass of the generation time distribution and test positivity curve — we use $L = 30$ in PMMH when fitting to REACT-1 data, again increasing this to $L = 50$ or $L = 70$ when estimating the marginal posterior distribution of the hidden states.

#### 3 Eales approach knot-spacing and equivalence with the SIMPLE approach

In this section we fit five models to REACT-1 data from rounds 14-to-19 (134 observations over 204 days), showing step-by-step the transition from the Eales approach to a slightly modified version of the SIMPLE approach.

All four implementations of the Eales approach use 200 warm-up iterations and 300 sampling iterations. As the data length is not divisible by 5, a target knot spacing of 5 days results in an effective spacing of 4.95 days. All other parameters are set to their default values.

For the Eales approach without splines, we fit the following model using Stan:

$$b_t \sim \text{Normal}\left(2b_{t-1} - b_{t-2}, \sigma_{Eales}\right)$$

$$P_t = \text{logit}^{-1}(b_t)$$

$$n_t^+ \sim \text{Beta-binomial}\left(n_t, \alpha_t, \beta_t\right)$$

where $\alpha_t = P_t(1/\rho - 1)$ and $\beta_t = (1 - P_t)(1/\rho - 1)$. Uniform prior distributions are used for $b_1$, $b_2$, and $\rho$, and a weakly informative inverse-gamma prior distribution with shape and scale 0.0001 is used for $\sigma_{Eales}$. All source code is available on GitHub.

For the SIMPLE approach with a logit link function, we implement the following epidemic model:

$$\tilde{r}_t = \tilde{r}_{t-1} + \epsilon_t, \quad \epsilon_t \sim \text{Normal}\left(0, \sigma\right)$$

$$\text{logit}P_t = \text{logit}P_{t-1} + \tilde{r}_t$$

and retain the typical beta-binomial observation model. This model is equivalent to the Eales model without splines, expressed in a form better suited to SMC methods. To match the Eales model, we use the same inverse-gamma prior distribution for $\sigma$ and artificially extend the data by 3 days at the start and finish, assuming $n_t = 0$ on these days. Minor differences remain due to how initial values are handled.

The Eales approach with a binomial observation model and spline knots every 5 days took 1m 59s to fit. Replacing the binomial distribution with a beta-binomial distribution increased the fitting time to 5m 13s due to the additional computation required, although the convergence diagnostics (Rhat and ESS) improved as the beta-binomial model fits the data better. Decreasing the knot-spacing from 5 days to 1 day further increased the fitting time to 8m 45s. Finally, fitting the Eales model without splines took a similar amount of time and produced identical results (allowing for Monte Carlo error). Fitting the equivalent model using SMC took 50s and produced nearly identical results. These results are summarised in Table S1.

The sole reason for placing knots more than one day apart is to reduce computational burden. As the splines are unnecessary when fitting on the same timescale as our observations (unless we wish to interpolate at a higher resolution), and the SIMPLE approach (on logit$P_t$) is equivalent to the daily Eales approach without splines, the "ideal" Eales approach is equivalent to the SIMPLE approach on logit$P_t$. As the SIMPLE approach is approximately 9x faster to fit (on this dataset), we recommend using the SIMPLE approach over the Eales approach in this scenario.

This difference is expected to be greater on longer datasets, as the SIMPLE approach scales better with the number of observations, although for slowly-varying epidemics where knots can be placed farther apart without biasing results, there may be utility in using the Eales approach with splines.

Table S1: Fitting diagnostics and parameter estimates for the binomial Eales approach with spline knots every 5 days, the beta-binomial Eales approach with spline knots every 5 days, the beta-binomial Eales approach with daily spline knots, the Eales approach without splines (a random walk on model coefficients), and a modified version of the SIMPLE approach modelling logit$P_t$.

| Approach | Eales | Eales | Eales | Eales (no splines) | SIMPLE (logit) |
|---|---|---|---|---|---|
| Obs. model | Binomial | Beta-binom | Beta-binom | Beta-binom | Beta-binom |
| Knot spacing (days) | 5 | 5 | 1 | 1 | NA |
| Duration | 1m 59s | 5m 13s | 8m 45s | 8m 57s | 50s |
| Max $\hat{R}$ | 1.06 | 1.02 | 1.04 | 1.03 | 1.01 |
| Min ESS | 59 | 230 | 273 | 210 | 143 |
| $\hat{\sigma}$ (5-day) | 0.22 | 0.16 | NA | NA | NA |
| | (0.14, 0.39) | (0.098, 0.25) | NA | NA | NA |
| $\hat{\sigma}$ (1-day) | NA | NA | 0.014 | 0.014 | 0.015 |
| | NA | NA | (0.0089, 0.023) | (0.0087, 0.023) | (0.0093, 0.022) |
| $\hat{\rho} \times 10^4$ | NA | 1.9 | 1.8 | 1.8 | 1.8 |
| | NA | (1.0, 3.2) | (0.83, 3.0) | (0.90, 3.2) | (0.71, 3.1) |

### 4 Comparisons with the original Eales approach

In this section we compare the original Eales approach with our modified version on the REACT-1 dataset.

The “original” approach in this section assumes a binomial observation distribution, sets the “max_treedepth” HMC hyperparameter to 10, and uses 1,000 warmup iterations and 19,000 sampling iterations, with the warmup and sampling iterations increased as needed to achieve convergence (Table S2).

The “modified” approach in this section matches the approach described in the main paper: a beta-binomial observation distribution is used, the “max_treedepth” HMC hyperparameter is increased to 15, and the number of warmup and sampling iterations is decreased to 200 and 300, respectively.

Both the original and modified approaches presented here have been updated from the source code to use new Stan syntax and cmdstanr (also necessitating updated processing code in R) and are initialised at plausible parameter values to reduce computation during initialisation. Our implementations of both the “original” and modified approaches are available on GitHub.

Runtime (measured on a 2021 M1 MacBook Pro) and convergence diagnostics are provided in Table S2. On shorter periods with lower overdispersion (study rounds 8-to-13 and 14-to-16), the original approach converged within the default number of iterations, taking 1m 51s to 2m 36s to fit, while the modified approach took 35s to 56s. On periods where overdispersion was greater (study rounds 1-to-7 and 17-to-19), additional iterations were required for the original approach to converge, taking up to 17m 2s to fit, while the modified approach took up to 2m 58s to fit (study rounds 1-to-7). We were unable to obtain convergence of the original approach on the full dataset, even after 250,000 iterations and 31 hours of computation, while the modified approach completed in 37m 42s.

Figure S3 presents results from fitting both models to the REACT-1 data by (grouped) study round. While both approaches produce similar results during rounds 8-to-13 and 14-to-16 (when overdispersion is estimated to be lower), there are substantial differences during rounds 1-to-7 and 17-to-19. The original approach produces estimates of $r_t$ and $P_t$ that are more variable, as any extra-binomial noise must be absorbed by variation in $r_t$. Furthermore, the posterior predictive credible intervals for observed swab positivity are wider under the modified approach (see Figure S4 for a closer look at rounds 1-to-7), due to the allowance for overdispersion.

Table S2: Runtime, convergence diagnostics, and coverage of 95% credible intervals for $n_t^+$ for the original and modified Eales approaches. In some cases, additional iterations were required to obtain convergence of the original model. 50,000 iterations (2,000 warmup + 48,000 sampling) were used for rounds 17-to-19 and 100,000 iterations (10,000 warmup + 90,000 sampling) were used for rounds 1-to-7. We attempted up to 250,000 iterations on the full dataset, however convergence was not achieved.

| **Round** | **All rounds** | **1-to-7** | **8-to-13** | **14-to-16** | **17-to-19** |
|---|---|---|---|---|---|
| Observations | 400 | 147 | 119 | 67 | 67 |
| Duration (days) | 700 | 217 | 195 | 100 | 86 |
| **Original approach** (1,000 warmup + 19,000 sampling) | | | | | |
| Runtime | 13m 12s | 3m 27s | 2m 36s | 1m 51s | 1m 49s |
| Max $\hat{R}$ | 3.02 | 1.39 | 1.01 | 1.00 | 1.12 |
| Min ESS | 3 | 8 | 577 | 648 | 18 |
| Converged? | No | No | Yes | Yes | No |
| Coverage | - | - | 94.1% | 91.0% | - |
| **Original approach** (extra samples, see caption) | | | | | |
| Runtime | 31h 4m 55s* | 17m 2s | - | - | 4m 29s |
| Max $\hat{R}$ | 3.62 | 1.05 | - | - | 1.02 |
| Min ESS | 2 | 74 | - | - | 122 |
| Converged? | No | Weakly | - | - | Yes |
| Coverage | - | 89.1% | - | - | 94.0% |
| **Modified approach** (200 warmup + 300 sampling) | | | | | |
| Runtime | 38m 54s** | 2m 58s | 1m 14s | 43s | 35s |
| Max $\hat{R}$ | 1.04** | 1.02 | 1.02 | 1.01 | 1.01 |
| Min ESS | 152** | 380 | 259 | 245 | 260 |
| Converged? | Yes | Yes | Yes | Yes | Yes |
| Coverage | 96.5% | 96.6% | 97.5% | 97.0% | 98.5% |

*This was timed on the Imperial College HPC cluster so the runtime is not directly comparable with the other approaches. **In order to obtain convergence of the modified approach on the full REACT-1 dataset, the "maximum tree-depth" HMC hyperparameter was increased from 15 to 16.

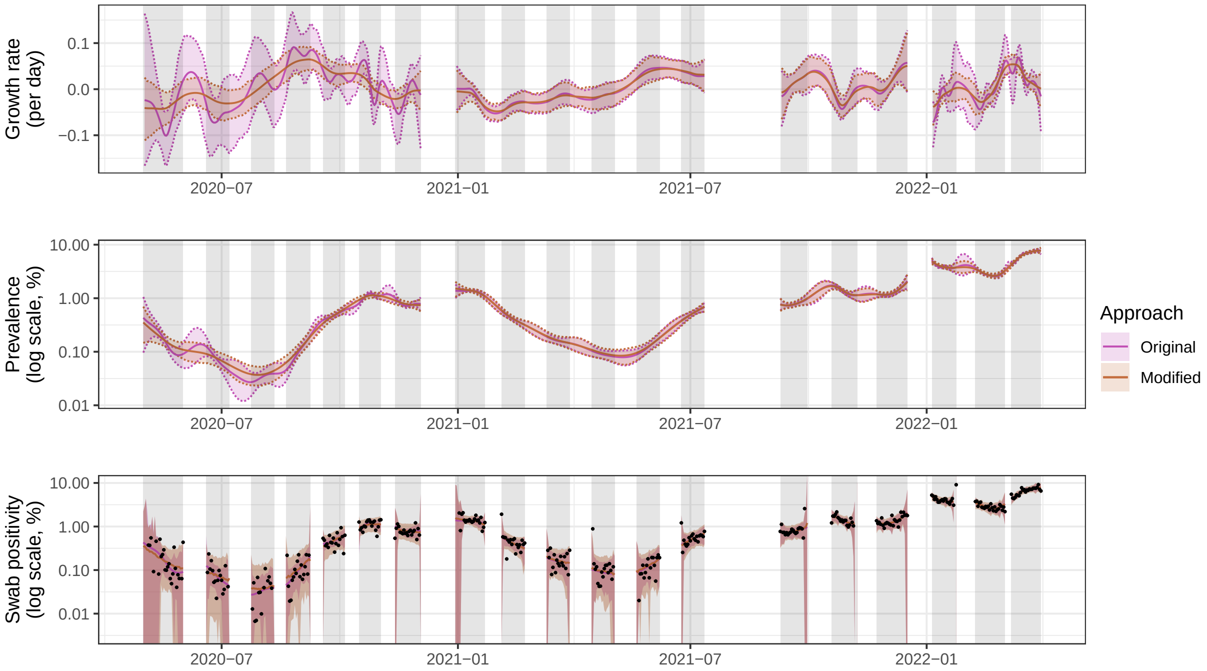

Figure S3: Model results from fitting the original and modified Eales approaches to the REACT-1 dataset, grouped by study rounds 1-to-7, 8-to-13, 14-to-16, and 17-to-19. Solid lines show the posterior mean of each quantity, while shaded regions and dashed lines show 95% credible intervals. Gray shading indicates the periods when the data were collected.

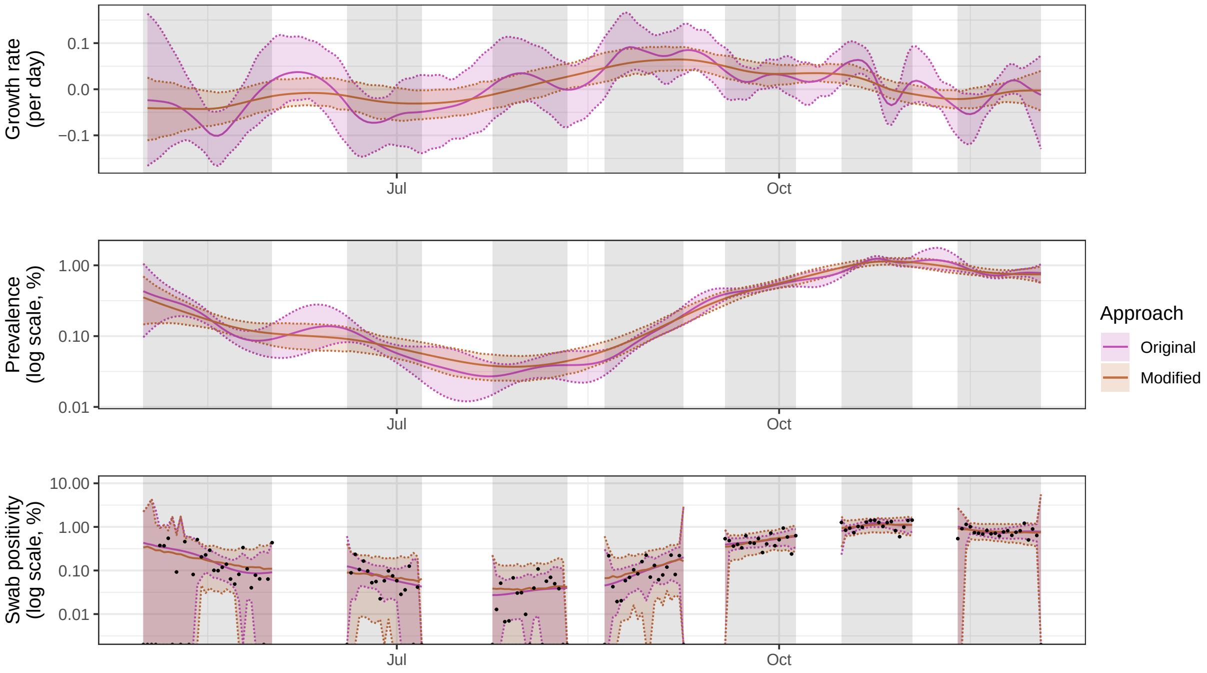

Figure S4: Model results from fitting the original and modified Eales approaches to rounds 1-to-7 of the REACT-1 dataset. Solid lines show the posterior mean of each quantity, while shaded regions and dashed lines show 95% credible intervals. Gray shading indicates the periods when the data were collected.

#### 5 RT-PCR test sensitivity curves for SARS-CoV-2

The Abbott approach relies upon an externally-estimated infection-to-swab-positivity curve that describes how likely a given individual is to return a positive test result at a given time from infection. The use of this curve allows us to estimate infection *incidence* (and thus quantities such as the reproduction number) from prevalence data while accounting for the sensitivity of the test. By modelling swab positivity as a convolution of past incidence with this curve, we are also imposing additional smoothness assumptions on the observed data: even if the underlying incidence is not smooth, the observed prevalence will be.

By default, the Abbott approach uses a sensitivity curve estimated by Hellewell, Russell, Matthews, et al. [3], which reflects the sensitivity of self-administered nasopharyngeal RT-PCR tests by healthcare workers for SARS-CoV-2 in England in early 2020. A total of 241 PCR tests from 27 individuals were used to estimate this curve. Their median curve suggests a peak sensitivity of 78.5% at 4 days from infection and declines to a sensitivity of less than 10% at 20 days from infection.

Other estimates of time-varying RT-PCR sensitivity for SARS-CoV-2 are available and highlight considerable uncertainty in the literature. Binny, Priest, French, et al. [4] used a total of 12,501 tests from 4,196 individuals that were tested using healthcare professional-administered nasopharyngeal RT-PCR tests for SARS-CoV-2 between June 2020 and November 2021. They found a peak sensitivity of 92.7% at 5 days from infection, declining to a sensitivity of less than 10% after 38 days. Kucirka, Lauer, Laeyendecker, Boon, and Lessler [5] pooled data from seven studies (a total of 1,330 swabs) to estimate a sensitivity curve peaking at 80.9% after 8 days, although their curve stops on day 21 at a sensitivity of 37%. These curves are presented in Figure S5.

Substituting the Hellewell et al. (2021) curve for the Binny et al. (2023) curve and refitting the model to the REACT-1 dataset produces Figure S6. While swab positivity estimates are largely similar, there are differences in the estimated growth rates. We also show the estimated infection incidence, demonstrating the substantial effect that the choice of sensitivity curve has on this quantity.

In the main paper, we hypothesised that the Abbott approach occasionally undercovers the growth rate due to additional smoothing assumptions imposed by the PCR sensitivity curve. As the Binny et al. (2023) curve represents a convolution over a longer time-period, it enforces additional smoothing on this quantity. We reproduce Figure 4 from the main paper in Figure

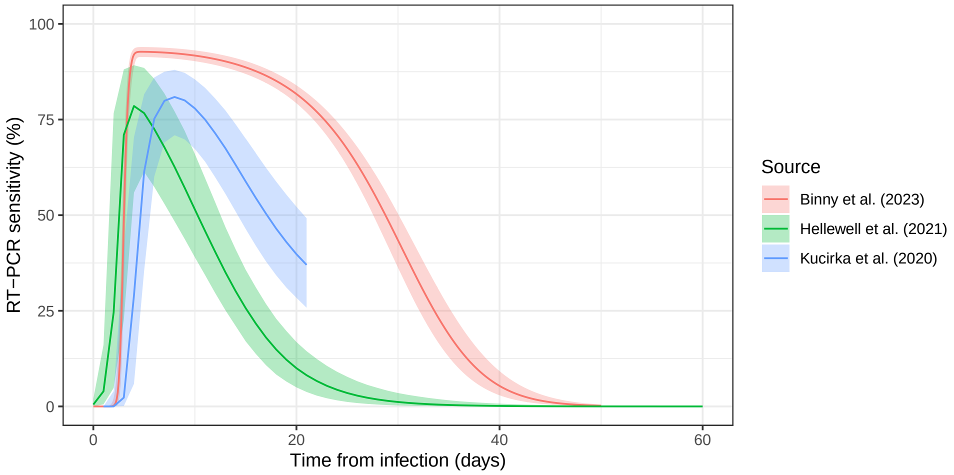

Figure S5: Median estimated RT-PCR sensitivity for SARS-CoV-2 and corresponding 95% credible intervals from three different sources, highlighting substantial differences in the literature. The Abbott approach uses curves from Hellewell et al. (2021) [3].

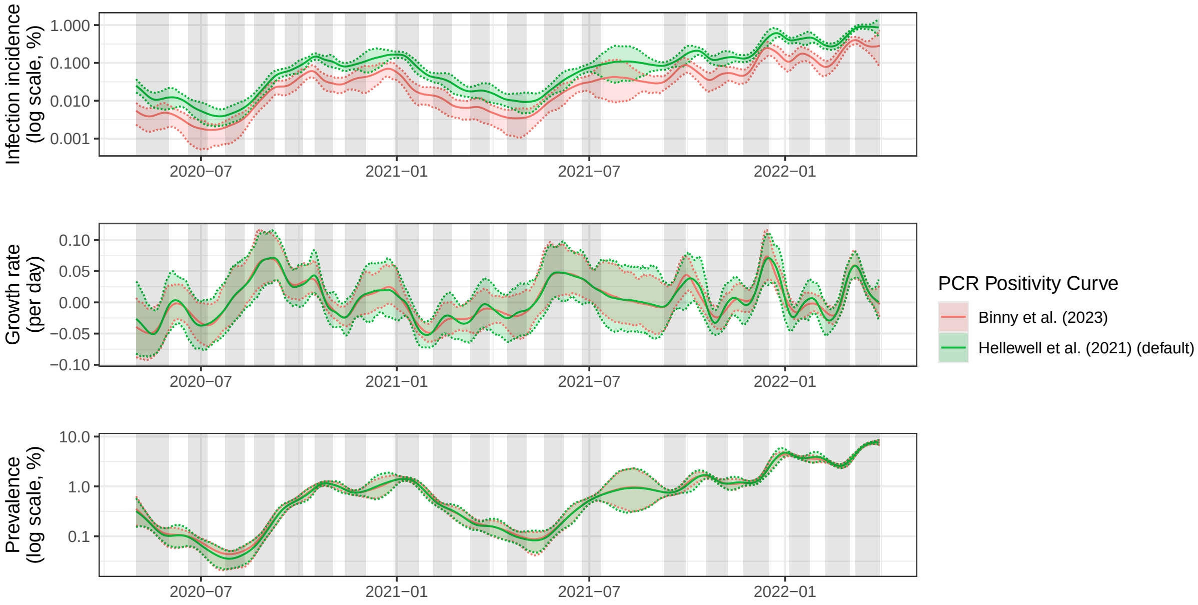

Figure S6: Results from fitting the Abbott approach to REACT-1 data using two different PCR positivity curves.

S7, additionally fitting the Abbott approach with the Binny et al. (2023) curve, and find that this hypothesis is supported. The Binny et al. (2023) curve results in even more smoothing of the growth rate estimates and also results in a decrease in accuracy of the estimated prevalence (when fit to simulations from the SIMPLE, Eales, and Abbott (Hellewell) approaches).

The SIMPLE approach also relies upon an externally-estimated infection-to-swab-positivity curve when estimating the reproduction number $R_t$. In Figure S8, we compare estimates by applying this approach to the REACT-1 dataset using the default Hellewell et al. (2021) curve alongside the Binny et al. (2023) curve. The latter curve results in a greater estimated value of

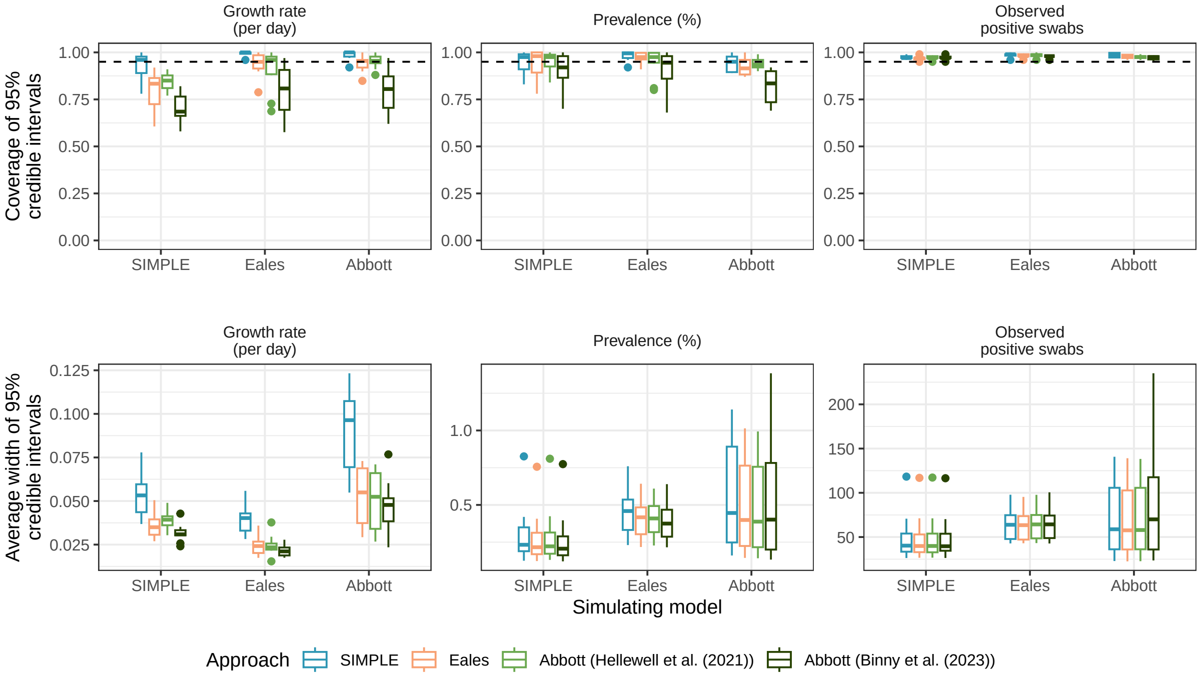

Figure S7: Coverage and average width of 95% credible intervals for the growth rate, prevalence, and observed swab positivity from fitting the SIMPLE (extra-binomial), Eales, and Abbott approaches (with two different PCR positivity curves) to 10 simulated datasets from each model. Boxes present the interquartile range of the results with the median shown as a horizontal line. Whiskers extend to the most extreme data point within 1.5 times the interquartile range from the box. Outliers are shown as points.

$\sigma_R$ of 0.10 (0.068, 0.15) (the central estimate using the Hellewell curve is 0.069 (0.050, 0.097)), which is a result of the additional smoothing of incidence imposed by the Binny curve (thus $R_t$ must become more variable to fit the same data). This greater value of $\sigma_R$, alongside the additional smoothing of incidence imposed by the Binny curve, means $R_t$ fluctuates more, resulting in substantially wider credible intervals for $R_t$.

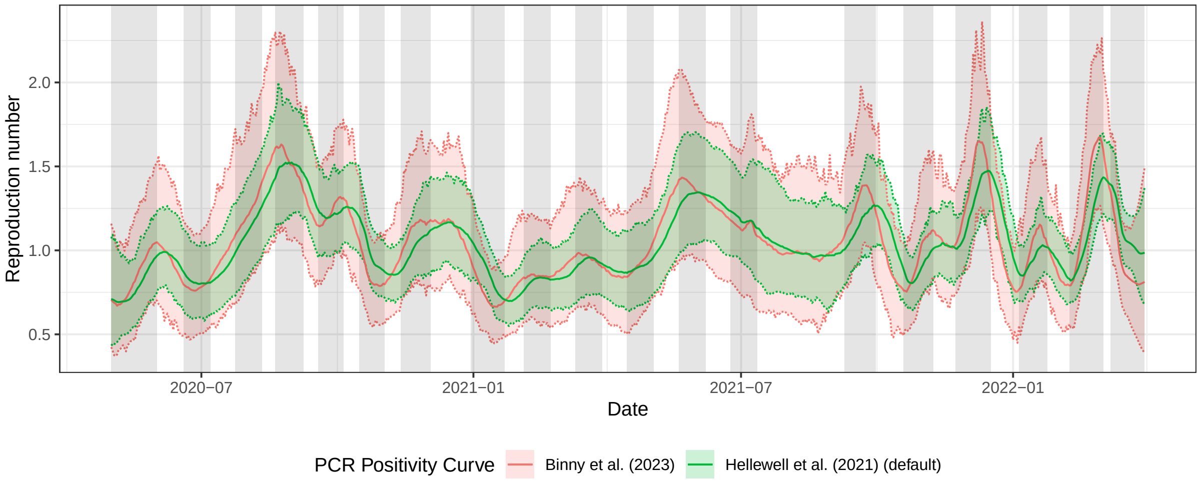

Figure S8: Results from fitting the SIMPLE approach for $R_t$ to REACT-1 data using two different PCR positivity curves. Solid lines show the posterior mean of each quantity, while shaded regions and dashed lines show 95% credible intervals. Gray shading indicates the periods when the data were collected.

### 6 REACT-1 survey weights

#### 6.1 Observed survey weights

We claim in the main text that REACT-1 survey weights are well modelled by log-normal distributions with scale parameters ranging between 0.392 and 0.645. We present histograms of the observed survey weights from each study round, along with fitted log-normal distributions (using maximum likelihood estimation) in Figure S9. The fitted distributions are generally in agreement with the data, although the observed weights are more centred around the mode than suggested by the fitted distributions. The "sdlog" parameter included in sub-figure headings is the scale parameter of the fitted log-normal distribution, which is equivalent to the parameter $\xi$ that we use when generating synthetic weighted data.

#### 6.2 Survey weights and observed swab positivity

While we do not explicitly model the relationship between survey weights and observed swab positivity when fitting the model to data, it is informative to investigate this relationship empirically. Specifically, we fit the following logistic regression model to REACT-1 data using the *glm* function in R:

$$\log \frac{P_i}{1 - P_i} = \beta_0 + \beta_1 \log w_i$$

where $P_i$ is the probability that the $i^{th}$ swab, with survey weight $w_i$, is positive.

Very crudely, if we assume that $P_i = kw_i^q$ and that $P_i$ is small, then we can show that $\beta_0 \approx q \log k$ and $\beta_1 \approx q$. Thus, the $\beta_1$ coefficient tells us about the relationship between survey weights and observed swab positivity. Specifically, $\beta_1 > 0$ implies those with greater weights (lower response probability) are more likely to test positive, and $\beta_1 = 1$ implies a linear relationship (like the one we assume for simulated weighted data).

Table S3 presents summary statistics of observed REACT-1 survey weights by round, and the $\beta_1$ coefficient (and associated p-value) from this logistic regression model. In 14 out of 19 study rounds, the $\beta_1$ coefficient is statistically significantly greater than 0 at the $\alpha = 0.05$ significance level, suggesting that those with greater survey weights (i.e., people with characteristics associated with lower response probabilities) are more likely to test positive, although the estimated value is always less than 1, suggesting a sublinear relationship. This relationship also appears to break down in study rounds 18 and 19, rounds in which the population prevalence was greatest and incentives to improve response rates were introduced.

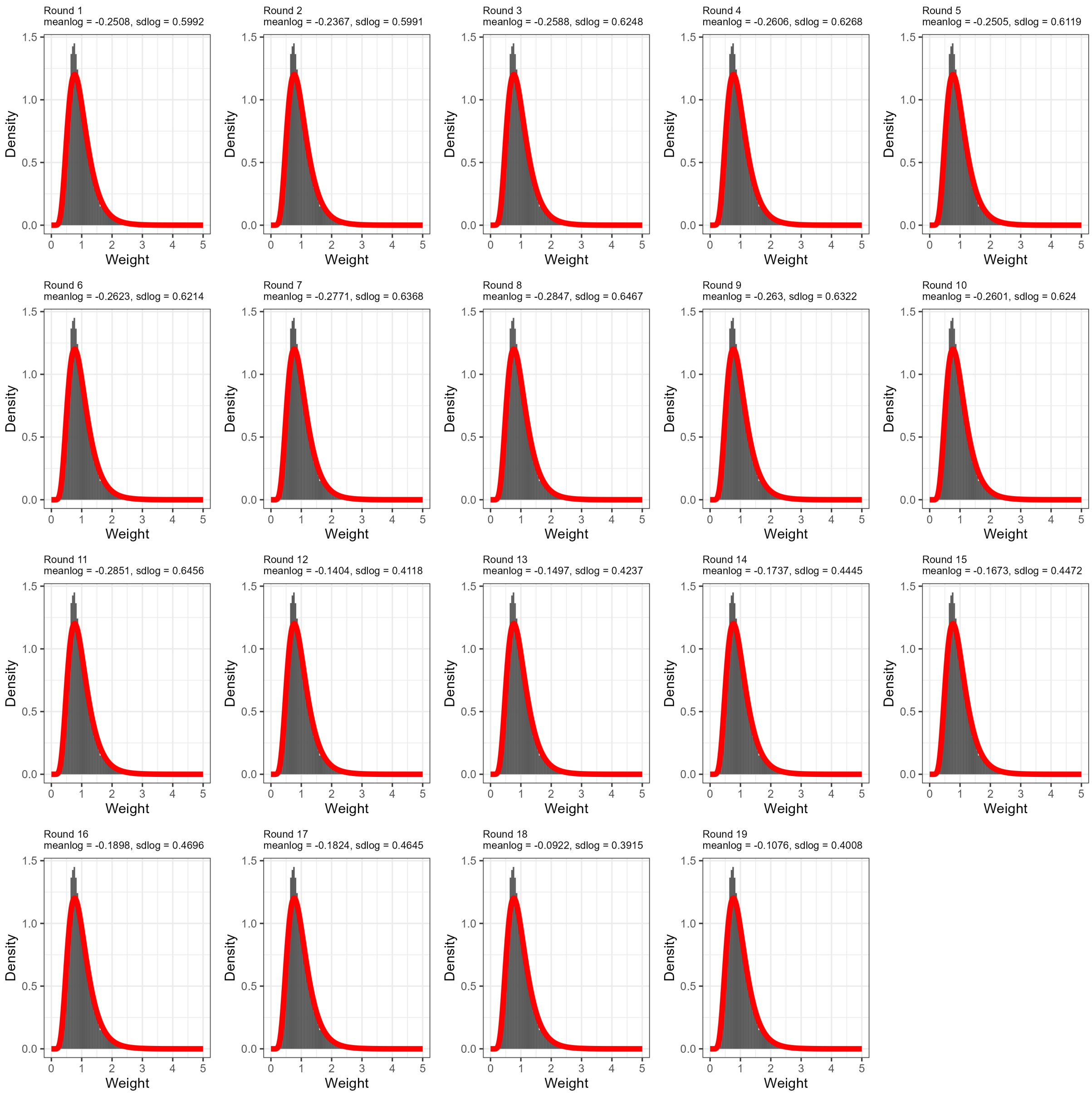

Figure S9: Histogram of REACT-1 survey weights by study round (grey bars) and fitted (by maximum likelihood estimation) log-normal distributions (red lines). The parameters of the fitted distributions are given in the subheadings of each panel.

Table S3: Summary statistics and logistic regression coefficient (see text) for observed survey weights in the REACT-1 study.

| Round | Mean weight | Median weight (2.5th quantile, 97.5th quantile) | Coefficient (p-value) |
|---|---|---|---|
| 1 | 0.953 | 0.724 (0.284, 2.96) | 0.307 (p = 0.017) |
| 2 | 0.964 | 0.736 (0.287, 2.88) | 0.246 (p = 0.089) |
| 3 | 0.96 | 0.718 (0.269, 3.03) | 0.372 (p = 0.069) |
| 4 | 0.961 | 0.717 (0.271, 3.07) | 0.610 (p <0.001) |
| 5 | 0.96 | 0.723 (0.282, 3.01) | 0.408 (p <0.001) |
| 6 | 0.958 | 0.714 (0.276, 3.09) | 0.298 (p <0.001) |
| 7 | 0.956 | 0.697 (0.273, 3.16) | 0.361 (p <0.001) |
| 8 | 0.954 | 0.693 (0.266, 3.19) | 0.299 (p <0.001) |
| 9 | 0.962 | 0.711 (0.266, 3.08) | 0.324 (p <0.001) |
| 10 | 0.959 | 0.719 (0.269, 3.02) | 0.438 (p <0.001) |
| 11 | 0.949 | 0.702 (0.253, 3.09) | 0.221 (p = 0.112) |
| 12 | 0.952 | 0.830 (0.440, 2.19) | 0.762 (p <0.001) |
| 13 | 0.949 | 0.820 (0.430, 2.23) | 0.696 (p <0.001) |
| 14 | 0.938 | 0.794 (0.408, 2.32) | 0.446 (p <0.001) |
| 15 | 0.944 | 0.797 (0.416, 2.32) | 0.567 (p <0.001) |
| 16 | 0.937 | 0.777 (0.391, 2.42) | 0.618 (p <0.001) |
| 17 | 0.939 | 0.786 (0.394, 2.39) | 0.400 (p <0.001) |
| 18 | 0.987 | 0.899 (0.426, 2.09) | -0.0357 (p = 0.473) |
| 19 | 0.978 | 0.869 (0.425, 2.19) | 0.0199 (p = 0.520) |

### 7 Supplementary results (simulated)

#### 7.1 Survey design and epidemic dynamics: hidden-state estimation

In the main paper, we considered the coverage and width of the 95% credible intervals for prevalence $P_t$ when fit to data from each model. We reproduce these results for the estimated growth rate $r_t$ in Figure S10. Unlike $P_t$, fitting non-weighted models to weighted data does not impact estimates of the growth rate $r_t$, although this assumes there are no temporal biases in survey weights. The basic model still struggles to estimate $r_t$ when the data are overdispersed, however, as the estimated $r_t$ must fluctuate significantly to account for observed noise in the data.

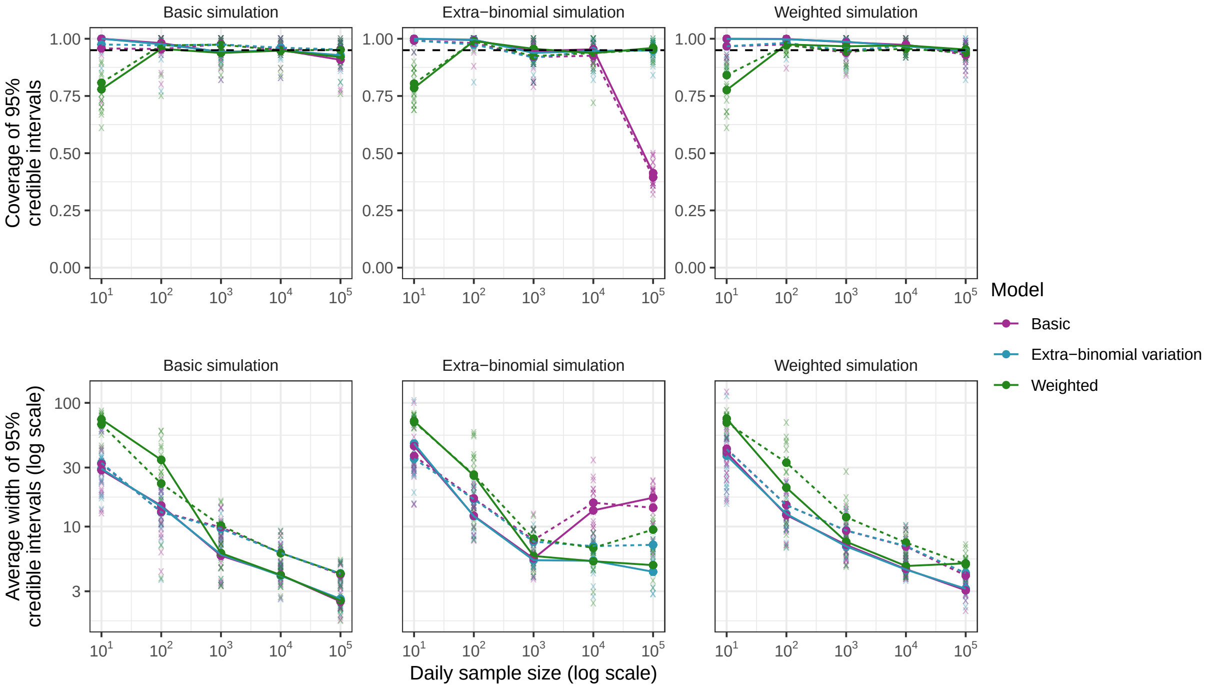

Figure S10: Coverage and average width of 95% credible intervals for the estimated growth rate $r_t$ from fitting all three models (basic: purple, extra-binomial: blue, weighted: green) to simulated data from each model (basic: column 1, extra-binomial: column 2, weighted: column 3). Results from individual simulations are shown as transparent crosses, with averages over 10 simulations shown as points connected by solid lines (for assumed $\sigma = 0.008$) and dashed lines (for assumed $\sigma = 0.016$). A range of assumed daily sample sizes $n_t$ are considered (x-axis). The horizontal black dashed line indicates the target coverage of 95%.

#### 7.2 Survey design and epidemic dynamics: parameter estimation

In addition to coverage and credible interval width for the hidden-states, we also consider how the estimation of model parameters varies with survey design and epidemic dynamics. Figure S11 presents the estimated value of $\sigma$, $\rho$ (where applicable) and $c$ (where applicable). As expected, higher $n_t$ results in more precise estimates of model parameters. All three models produce good estimates of $\sigma$, even when there is no extra-binomial variation or survey weight bias, suggesting there is little cost to including the additional parameter. When fitting the basic model to data featuring extra-binomial variation, the estimated value of $\sigma$ increases to compensate for the additional noise, with a greater effect observed at higher values of $n_t$. When fitting the weighted model to data featuring extra-binomial variation, the variance adjustment factor $c$ increases to account for this, so the weighted model still produces good estimates of $\sigma$.

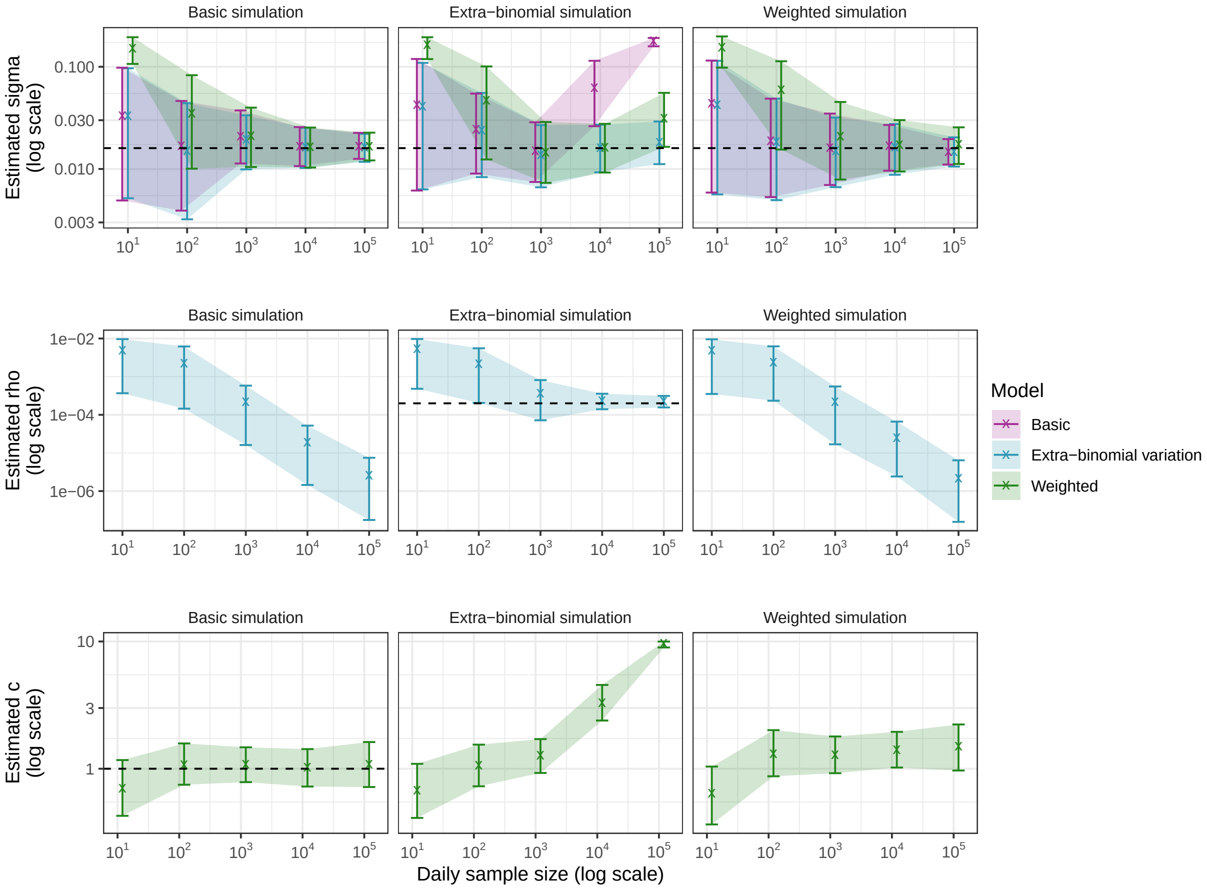

Figure S11: 95% credible intervals for model parameters from fitting all three models (basic: purple, extra-binomial: blue, weighted: green) to simulated data from each model (basic: column 1, extra-binomial, column 3, weighted: column 3) at varying values of $n_t$ and $\sigma = 0.016$. Black dashed lines denote true parameter values where applicable.

#### 8 Comparing model runtimes

Runtimes reported in the main paper reflect a single successful run of each model. However, runtimes can vary substantially across runs, even for the same model and dataset. To assess relative performance more robustly, we repeat each fitting procedure multiple times.

Observed runtimes depend on many factors, including dataset length, model complexity, parameterisation, model fit, and the system used to fit the model. The results in this section are based on subsets of the REACT-1 dataset and may not generalise to other settings. We use real data to avoid artefacts from fitting models to their own simulations, but caution that these comparisons remain context-specific.

We test each model on multiple subsets of the REACT-1 dataset of increasing length, always starting from study round 1 (1 May 2020). Each subset is defined by its final study round—rounds 1 through 13 in sequence, followed by rounds 15, 17, and 19 to reduce computation. Most models are fit 10 times per subset, except for the SIMPLE approach (reproduction number epidemic model) and the Eales approach, which are run 5 times each for rounds 13, 15, and 17, and 3 times for round 19.

All models use their default parameterisations, except for the Eales approach on all 19 rounds, where the HMC "max_treedepth" is increased to 16 to ensure convergence. For the SIMPLE approach with the reproduction number epidemic model, we use $N = 2000$ particles per PMMH likelihood evaluation for datasets ending at round 13 or later.

We present two runtime metrics: the time taken to complete a single successful fit (Figure S12, upper panel), and the average time taken to achieve convergence (Figure S12, lower panel). The latter accounts for failed fits, so is always equal to or greater than the former. As convergence is built into the stopping criteria for the SIMPLE approaches, only the Eales and Abbott approaches are affected by failed fits.

Stan models may fail to converge for two main reasons: (1) too few iterations, and (2) divergent chains. The first occurs when the data are too short (providing limited information) or too long (introducing many parameters), and affects both Stan-based models. The second (divergent chains) can occur regardless of dataset length and primarily affects the Abbott model. Improved parameter initialisation may reduce the frequency of these divergent chains, but is context-dependent and not explored further here.

The SIMPLE approach (growth rate epidemic model) and Abbott approach have comparable runtimes when successful, though the non-zero failure rate of the Abbott approach increases its average time per successful fit. The SIMPLE approach (reproduction number epidemic model) and the Eales approach are substantially slower overall. While runtimes for these two approaches are similar on shorter datasets, the SIMPLE approach (reproduction number epidemic model) is faster than the Eales approach on the longest dataset.

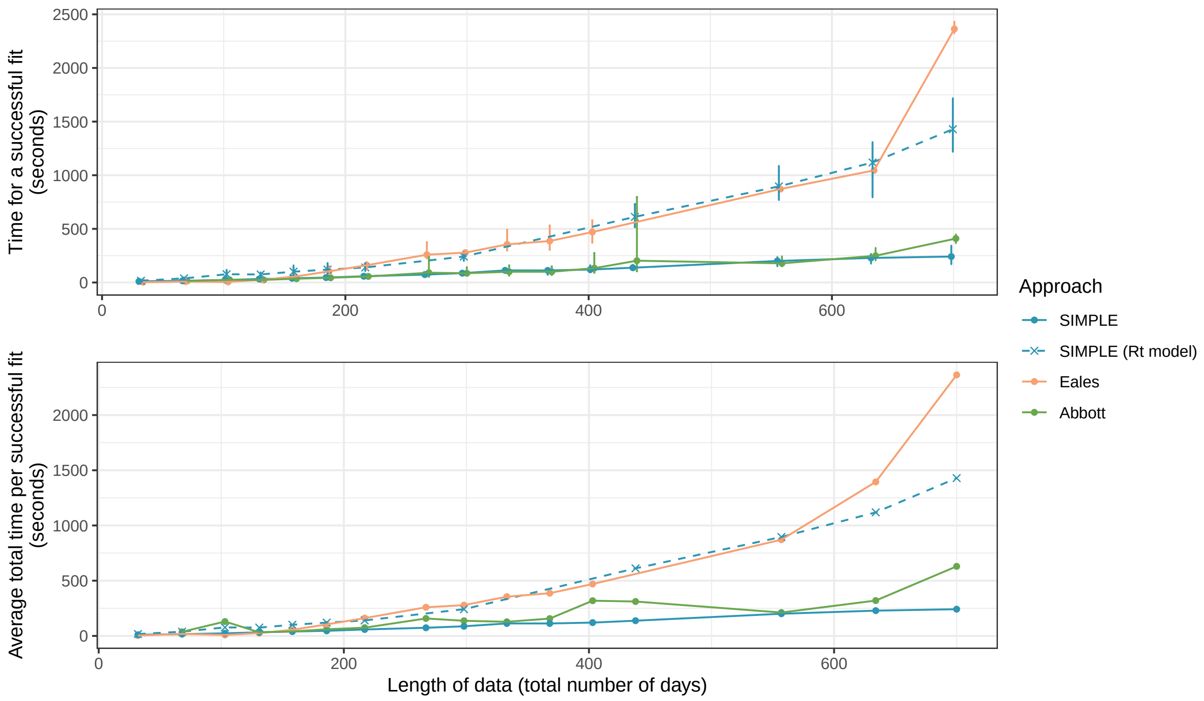

Figure S12: Runtimes for fitting each model to subsets of the REACT-1 dataset of increasing length (always starting from study round 1). The upper panel shows the average time taken to complete a single successful fit (points), with error bars showing the observed range (minimum to maximum). The lower panel shows the average time taken to achieve convergence, which is equal to or greater than the time for a single fit due to occasional failures. The Abbott approach failed to converge on the shortest dataset (round 1 only, 32 days), though this could likely be resolved by adjusting hyperparameters.

### 9 Supplementary results (REACT-1)

#### 9.1 Full study versus separate models

In the main paper, we present estimates of $r_t$, $P_t$, and $n_t^+/n_t$ from fitting to all 19 REACT-1 study rounds simultaneously (Figure 5), despite substantial variation in parameter estimates when fitting to shorter time periods (Table 3). In Figure S13, we compare hidden-state estimates from the full model with those obtained by fitting to shorter periods. Estimates are similar across rounds 1-to-7 and 8-to-13, although slightly wider credible intervals for $r_t$ are observed in rounds 14-to-16 and 17-to-19. These reflect greater uncertainty about model parameters and higher estimated values of $\sigma$.

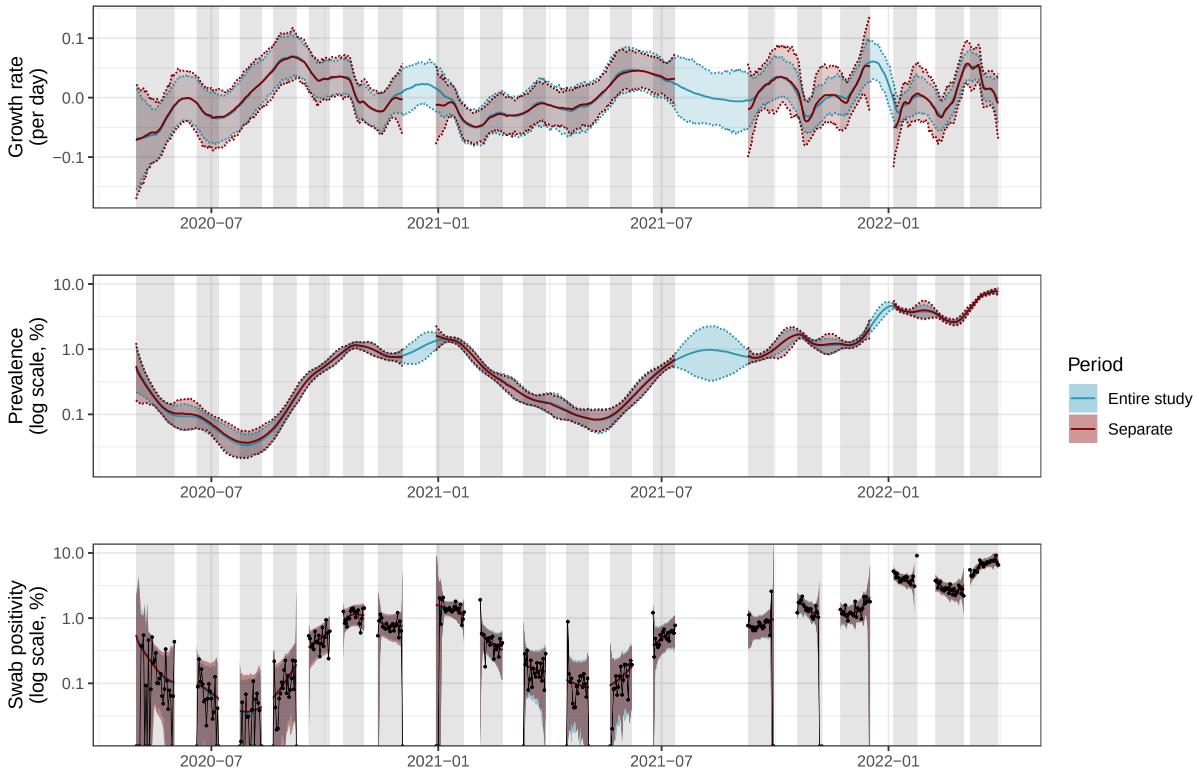

Figure S13: Estimates of the growth rate $r_t$, prevalence $P_t$, and observed swab positivity $n_t^+/n_t$ for SARS-CoV-2 in England between 1 May 2020 and 31 March 2022, based on data from the REACT-1 study. Blue curves present estimates from fitting to all 19 study rounds simultaneously while dark red curves present estimates from fitting to four shorter periods. Solid coloured lines show central estimates while shading and dashed lines show 95% credible intervals. Daily true observed swab positivity $n_t^+/n_t$ is shown in black points.

#### 9.2 Overdispersed versus basic model

We present estimates of $r_t$, $P_t$, and $n_t^+/n_t$ from the REACT-1 study using the beta-binomial model, allowing for extra-binomial variation in the data. We compare these estimates to those from the basic model in Figure S14. While estimates in rounds 8-to-13 and 14-to-16 are similar between models, estimates in rounds 1-to-7 and 17-to-19 are noticeably different - these are the periods in which the estimated value of $\rho$ is higher (Table 3).

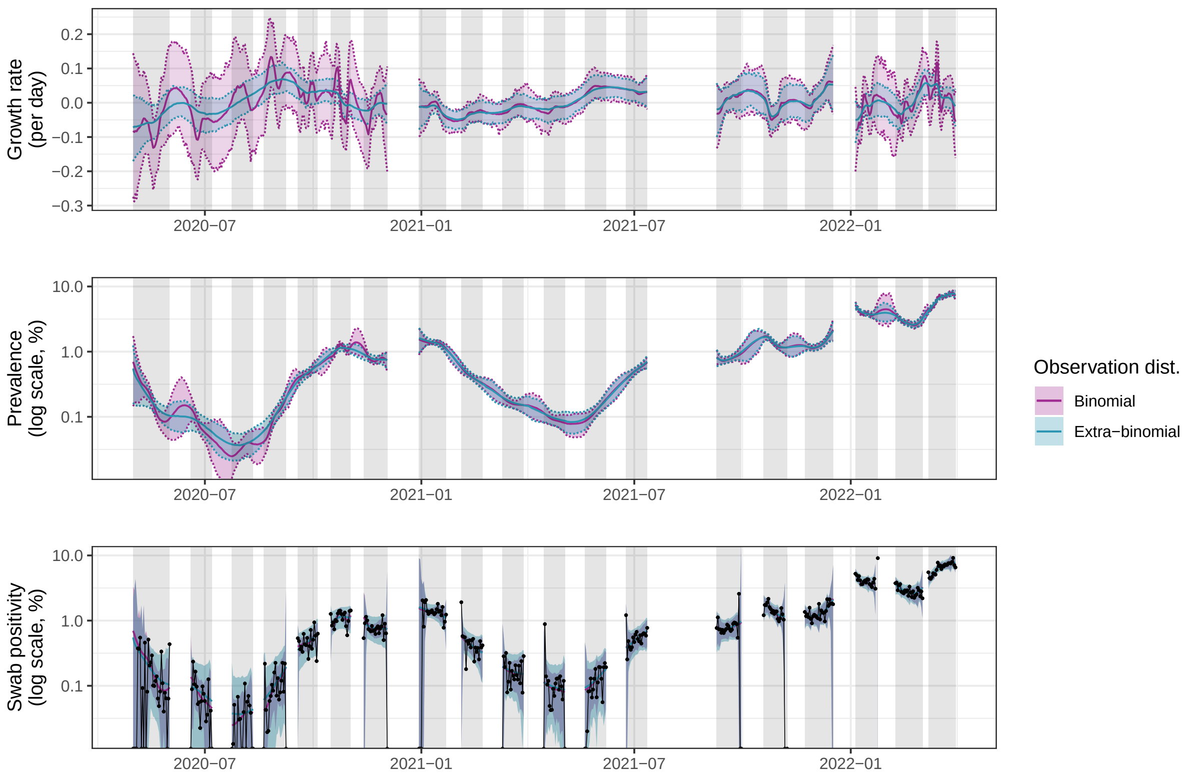

Figure S14: Estimates of the growth rate $r_t$, prevalence $P_t$, and observed swab positivity $n_t^+/n_t$ for SARS-CoV-2 in England between 1 May 2020 and 31 March 2022 using data from the REACT-1 study. Blue curves present estimates from the model allowing for extra-binomial variation while pink curves present estimates from the basic model. Solid coloured lines show central estimates while shading and dashed lines show 95% credible intervals. Daily true observed swab positivity $n_t^+/n_t$ is shown in black points.

#### 9.3 Weighted data

As survey weights applicable on a daily basis were not available for the REACT-1 study, we have thus far not fit the weighted model to these data. In Figure S15 we compare the extra-binomial model with the weighted model when fit to REACT-1 data, assuming that the provided survey weights (which are applicable on a round-by-round basis) are valid for use with daily data. Growth rate estimates are largely unchanged, although there are periods where population swab positivity is estimated to be greater in the weighted model.

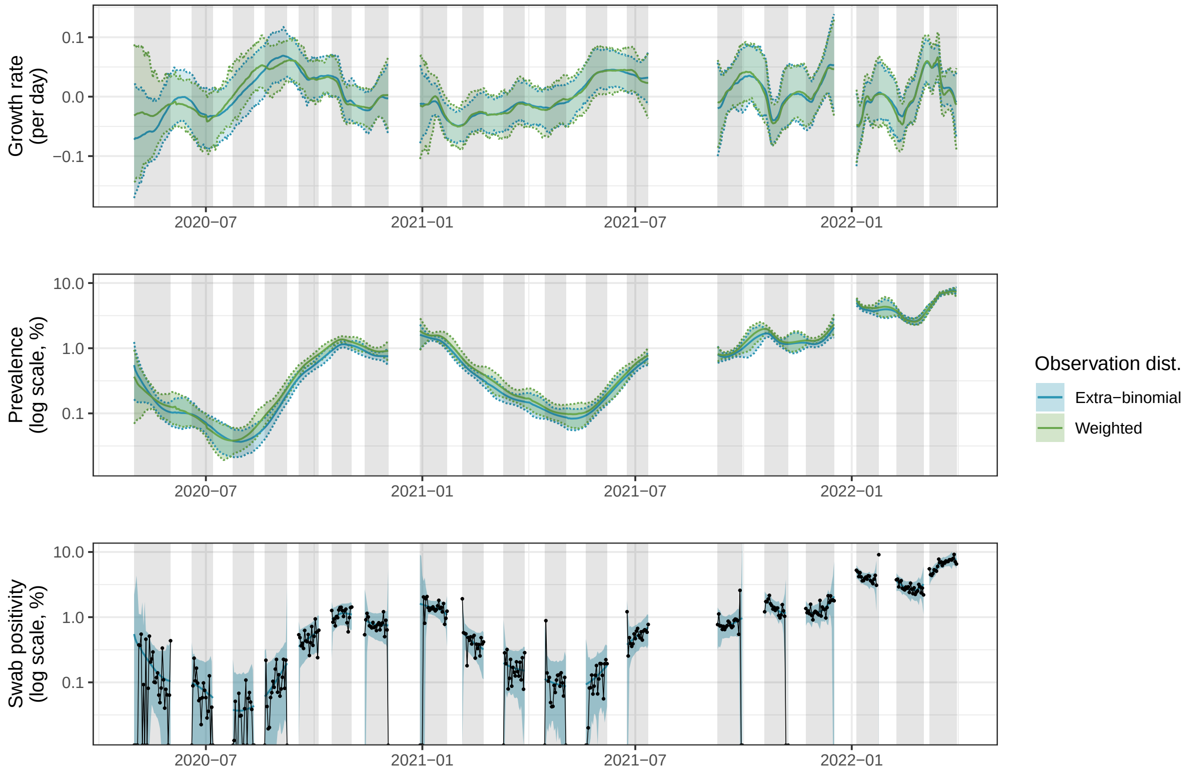

Figure S15: Estimates of the growth rate in swab positivity $r_t$, population swab positivity $P_t$, and observed swab positivity $n_t^+/n_t$ for SARS-CoV-2 in England between 1 May 2020 and 31 March 2022 using data from the REACT-1 study. Blue curves present estimates from the model allowing for extra-binomial variation while green curves present estimates from the weighted model, assuming that survey weights are valid on a day-by-day basis. Solid coloured lines show central estimates while shading and dashed lines show 95% credible intervals. Daily true observed swab positivity $n_t^+/n_t$ is shown in black points.

#### 10 Accounting for multiple variants

Genomic sequencing has become an increasingly important tool for monitoring the spread of infectious diseases. This was most notable during the COVID-19 pandemic, where the repeated emergence of new variants of concern necessitated the monitoring of variant-specific growth rates. Genomic sequencing of survey samples is time-consuming and expensive, so the estimation of variant-specific growth rates using sequencing from only a subset of samples is a key problem. We show how the SIMPLE approach can be extended to allow for this.

Specifically, using superscript $v = 1, \dots, V$ to denote variant, we define variant-specific growth rates and swab positivity:

$$r_t^{(v)} = r_{t-1}^{(v)} + \epsilon_t^{(v)}, \quad \epsilon_t^{(v)} \sim N(0, \sigma_r^{(v)})$$

$$P_t^{(v)} = P_{t-1}^{(v)} e^{r_t^{(v)}}$$

We retain the standard beta-binomial observation distribution for the number of total positive swabs, with overall prevalence defined as $P_t = \sum_v P_t^{(v)}$. We further assume that $n_t^s$ of the positive swabs are sent for genomic sequencing and that number of these swabs that are positive for each variant considered follows a multinomial distribution. Specifically:

$$n_t^+, \left\{ n_t^{(v)} \right\} \sim \text{Beta-binomial}(n_t, \alpha_t, \beta_t) \times \text{Multinomial} \left( n_t^s, \left\{ \frac{P_t^{(v)}}{P_t} \right\} \right)$$

where the "$\times$" operator denotes the product of two probability density functions. $\alpha_t = P_t(1/\rho - 1)$ and $\beta_t = (1 - P_t)(1/\rho - 1)$. We now have $V + 1$ parameters: $\sigma_r^{(v)}$, the standard deviation of the daily growth rate for each variant, and $\rho$, the overdispersion parameter.

The hidden-states associated with a given variant are initialised on the first day the variant is detected. We stop estimating the associated hidden-states at the end of the last round in which a given variant is detected.

We fit this model to REACT-1 data in two groups: rounds 8-to-13 (for Wildtype (WT), Alpha (AL), and Delta (DE) variants) and rounds 14-to-19 (for Delta, Omicron (OM), and Omicron BA.2 (BA) variants). Table S4 presents parameter estimates and 95% credible intervals. Figure S16 shows the estimated growth rates and prevalence for each variant. Crucially these estimates leverage data from both sequenced and unsequenced swabs to inform estimates of variant specific growth rates.

Table S4: Parameter estimates for the variant model fit to REACT-1 data.

| Rounds | $\sigma^{(WT)}$ | $\sigma^{(AL)}$ | $\sigma^{(DE)}$ | $\sigma^{(OM)}$ | $\sigma^{(BA)}$ | $\rho$ ($\times 10^4$) |
|---|---|---|---|---|---|---|
| 8-to-13 | 0.039 | 0.0077 | 0.014 | - | - | 1.3 |
| | (0.0014, 0.13) | (0.0013, 0.018) | (0.00089, 0.051) | - | - | (0.53, 2.3) |
| 14-to-19 | - | - | 0.016 | 0.035 | 0.020 | 1.7 |
| | - | - | (0.0084, 0.029) | (0.020, 0.062) | (0.007, 0.048) | (0.54, 3.3) |

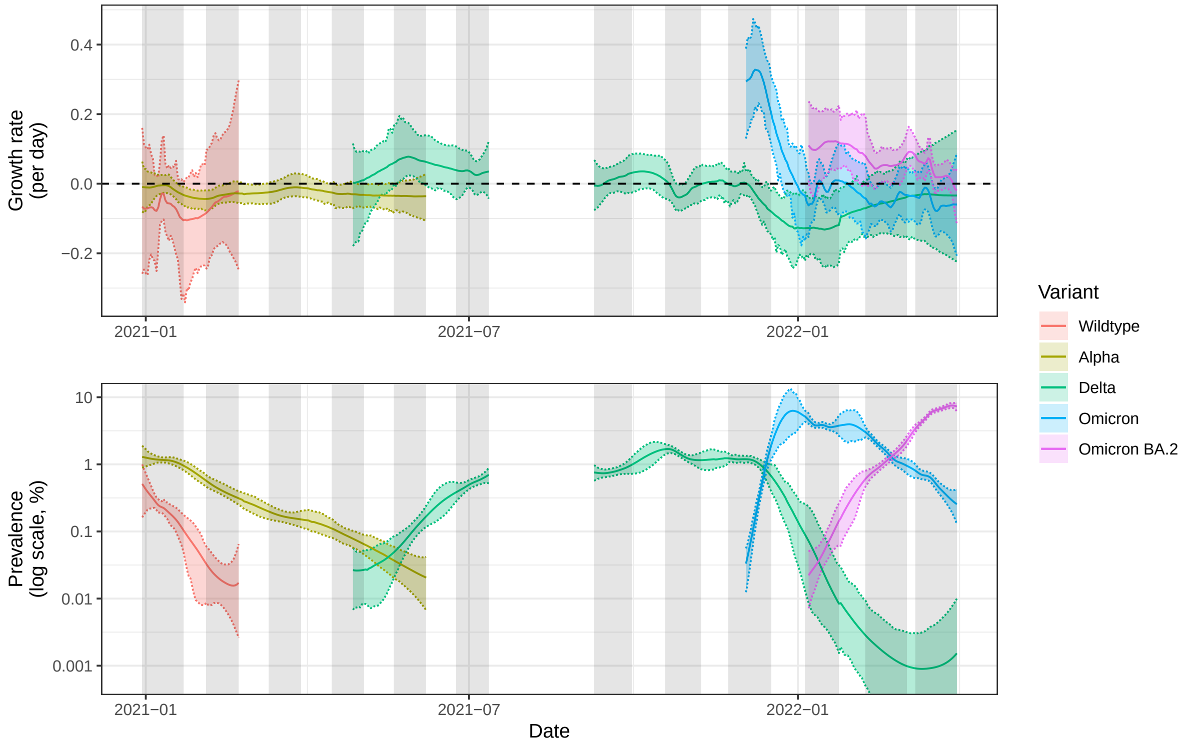

Figure S16: Estimated growth rates and prevalence of SARS-CoV-2 by variant between 30 December 2020 and 31 March 2022 using data collected in the REACT-1 study. Solid lines show the posterior mean of each quantity and shaded regions/coloured dashed lines show 95% credible intervals. Shaded regions indicate the periods when the data were collected. The horizontal black dashed line shows zero growth rate.

The observation model leveraged above can be derived by assuming that the probability of sequencing a given swab is independent of the variant, and that the total number of sequenced swabs is deterministic given the total number of positive swabs. Thus:

$$P\left(\{n_t^{(v)}\}, n_t^+, n_t^s | n_t, \cdots\right) = P\left(\{n_t^{(v)}\} | n_t^s, \cdots\right) P(n_t^s | n_t^+, \cdots) P(n_t^+ | n_t, \cdots)$$

where the first term is the multinomial distribution, the middle term is a Dirac delta at the observed $n_t^s$, and the final term is the beta-binomial distribution.

This framework readily allows for further analysis. Samples of growth rate advantages $(r_t^{v_a}/r_t^{v_b})$

can be obtained by comparing $r_t^{v,i}$ values across variants $v$, where $i$ indexes the particle number. Forecasts of variant prevalence can be obtained by projecting the model forward in time.

#### 11 Comparisons with UKHSA consensus estimates

During the COVID-19 pandemic, the UK Health Security Agency (UKHSA) produced consensus estimates of the growth rate and the reproduction number in England. These estimates were produced by combining estimates from a range of models that were fit to a variety of data sources (not including data from REACT-1) [6, 7]. The UKHSA estimates are the most official estimates of the growth rate and the reproduction number in England, so it is informative to compare our estimates with these.

UKHSA consensus estimates were reported, and are reported here, as the lower and upper bounds of the 90% credible intervals for the growth rate and the reproduction number. No central estimates were provided. Estimates were produced weekly and reflect data up to a few days before the estimate was produced (the exact gap varies by week). Figure S17 presents our estimates of the growth rate and the reproduction number from applying all approaches to the REACT-1 dataset, alongside the UKHSA estimates (shown in black).

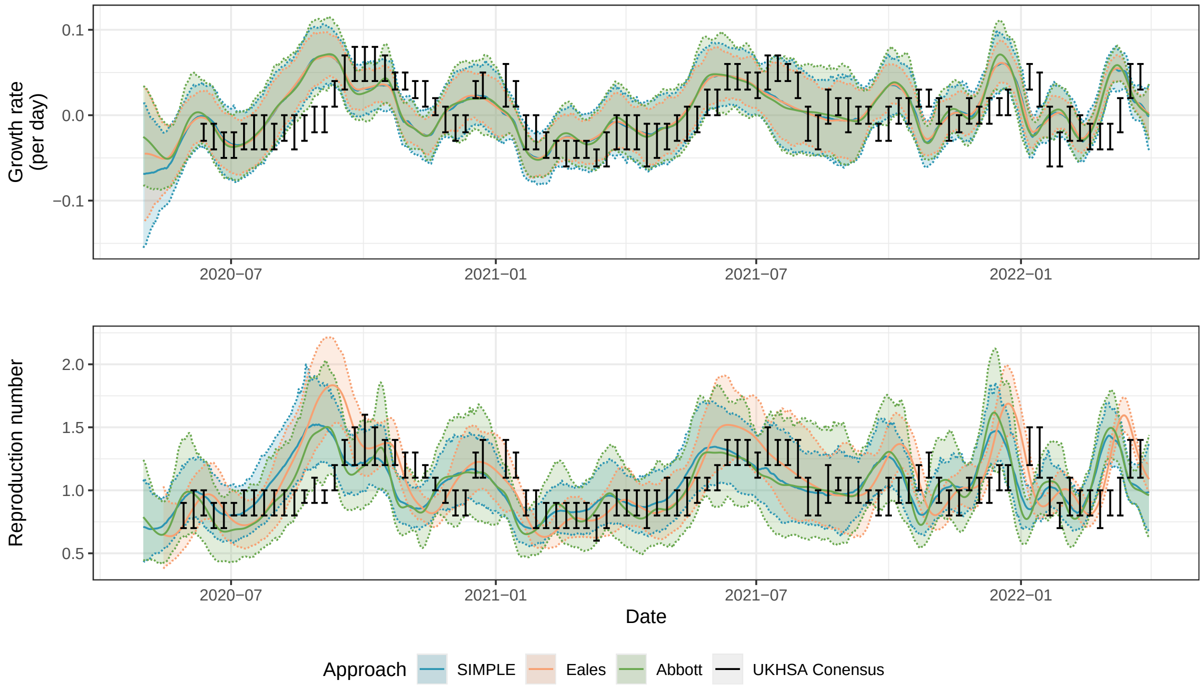

Figure S17: Comparison of our estimates of the growth rate and the reproduction number for SARS-CoV-2 in England with UKHSA consensus estimates. Our estimates are shown in blue (SIMPLE approach), orange (Eales approach), and green (Abbott approach). The UKHSA estimates are shown in black, with the lower and upper bounds of the 90% credible intervals shown as horizontal lines.

There are many reasons why our estimates differ from the UKHSA estimates:

1. The UKHSA estimates are consensus estimates produced by combining estimates from a

range of methods. This generally results in narrower credible intervals than those produced by any individual method, particularly where the individual models contributing to the consensus estimate agree. Consensus estimates can also result in more conservative estimates (closer to 0 for the growth rate and 1 for the reproduction number), as individual model variance is averaged out.

2. The UKHSA estimates are produced in real-time and are not updated retrospectively, whereas our approaches all use the full dataset to produce estimates. This explains why the UKHSA estimates lag behind our estimates: an increase in cases the following week can be explained by an increase in $r_t$ and $R_t$ today, but the UKHSA estimates will not reflect this until the increase is observed.

3. The UKHSA estimates are produced from different data sources to our estimates. The REACT-1 study did not contribute to the UKHSA estimates, which leveraged data such as reported cases and hospitalisations (among other, individual model-dependent, data sources).

4. All models make their own set of assumptions. In addition to serial interval and smoothing differences discussed in the main paper, the model structure can also impact estimates. Not all of the individual models in the UKHSA consensus estimates used the renewal model, for example.

5. The UKHSA report 90% credible intervals, while we report 95% credible intervals.

While consensus estimates like those produced by the UKHSA are generally more robust than estimates from any individual model, we are hesitant to use these comparisons as evidence that our approaches are producing inaccurate estimates. Points (2) and (3) above are particularly important: our estimates are produced retrospectively using high-quality data and are not subject to the same real-time constraints as the UKHSA estimates. Data that are available in real-time (such as reported cases and hospitalisations) are noisy and subject to biases which will impact all models that use them (and thus the consensus estimates). The instances where credible intervals do not overlap in Figure S17 largely occur due to delays in the UKHSA estimates, which would likely not exist if the estimates were produced using retrospective data.
